# Profiling SARS-CoV-2 mutation fingerprints that range from the viral pangenome to individual infection quasispecies

**DOI:** 10.1101/2020.11.02.20224816

**Authors:** Billy T. Lau, Dmitri Pavlichin, Anna C. Hooker, Alison Almeda, Giwon Shin, Jiamin Chen, Malaya K. Sahoo, ChunHong Huang, Benjamin A. Pinsky, HoJoon Lee, Hanlee P. Ji

## Abstract

**Background:** The genome of SARS-CoV-2 is susceptible to mutations during viral replication due to the errors generated by RNA-dependent RNA polymerases. These mutations enable the SARS-CoV-2 to evolve into new strains. Viral quasispecies emerge from *de novo* mutations that occur in individual patients. In combination, these sets of viral mutations provide distinct genetic fingerprints that reveal the patterns of transmission and have utility in contract tracing.

**Methods:** Leveraging thousands of sequenced SARS-CoV-2 genomes, we performed a viral pangenome analysis to identify conserved genomic sequences. We used a rapid and highly efficient computational approach that relies on k-mers, short tracts of sequence, instead of conventional sequence alignment. Using this method, we annotated viral mutation signatures that were associated with specific strains. Based on these highly conserved viral sequences, we developed a rapid and highly scalable targeted sequencing assay to identify mutations, detect quasispecies and identify mutation signatures from patients. These results were compared to the pangenome genetic fingerprints.

**Results:** We built a k-mer index for thousands of SARS-CoV-2 genomes and identified conserved genomics regions and landscape of mutations across thousands of virus genomes. We delineated mutation profiles spanning common genetic fingerprints (the combination of mutations in a viral assembly) and rare ones that occur in only small fraction of patients. We developed a targeted sequencing assay by selecting primers from the conserved viral genome regions to flank frequent mutations. Using a cohort of SARS-CoV-2 clinical samples, we identified genetic fingerprints consisting of strain-specific mutations seen across populations and de novo quasispecies mutations localized to individual infections. We compared the mutation profiles of viral samples undergoing analysis with the features of the pangenome.

**Conclusions:** We conducted an analysis for viral mutation profiles that provide the basis of genetic fingerprints. Our study linked pangenome analysis with targeted deep sequenced SARS-CoV-2 clinical samples. We identified quasispecies mutations occurring within individual patients, mutations demarcating dominant species and the prevalence of mutation signatures, of which a significant number were relatively unique. Analysis of these genetic fingerprints may provide a way of conducting molecular contact tracing.

## BACKGROUND

The source of the COVID-19 pandemic is the SARS-CoV-2 coronavirus, encoded by a single-stranded RNA molecule [1]. Given its airborne transmission, this virus has rapidly spread across disparate geographic regions and infected diverse populations [2]. An important genetic feature of the SARS-CoV-2 genome is the presence of viral mutations. There are two general categories of mutations based on their frequency among infected individuals **(Figure 1)**. Strain-level mutations are found in a relative high frequency among affected populations and provide viral genetic fingerprints from which one can trace the routes of transmission across broad geographic regions. This genetic information is useful in contact tracing for super-spreader events, where a given viral strain is introduced among a group of exposed individuals [3]. On a more granular scale, individual patients with active infections also show evidence of *de novo* mutations that occur as the virus replicates [4, 5]. These novel mutations define subclonal quasispecies that are private to an individual [6]. Occurring at a significantly lower population frequency, quasispecies-level mutations are frequently present in only a fraction of the total viral load of an infected individual and may be specific to a given patient. Sets of these mutations per a given virus provide personalized mutation profiles, in other words a distinct genetic fingerprint that is useful for characterizing transmission patterns of infection.

**Figure 1.**
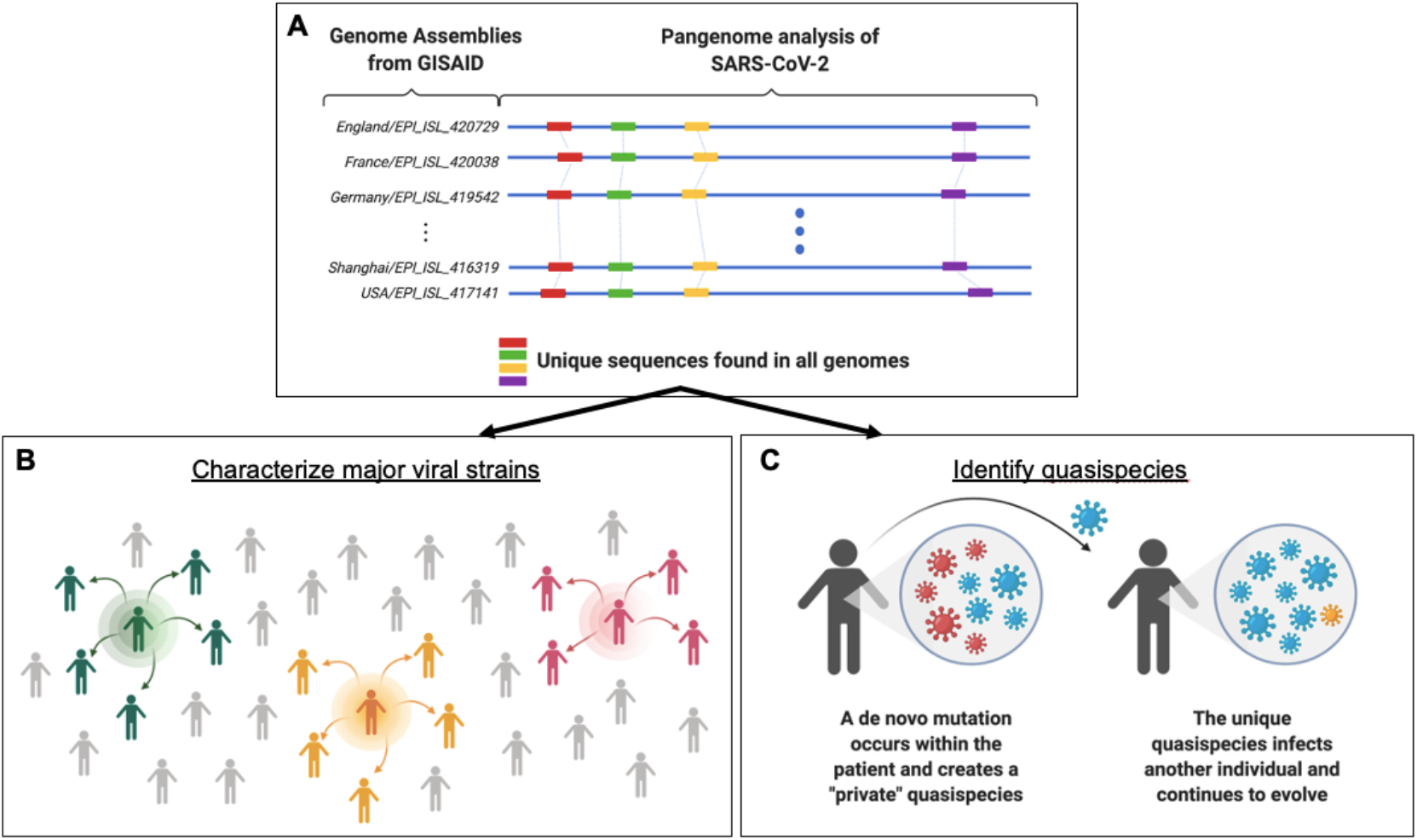
Framework for identifying population-level SARS-CoV-2 strains and lower frequency quasispecies through pangenome analysis. **(A)** GISAID currently has thousands of SARS-CoV-2 genomic sequences banked. By analyzing large numbers of viral genomes together (blue lines), one can pinpoint sequences that are present only once in a given genome but also occur consistently across all genomes (red, green, yellow and purple bars). These unique and conserved sequences can be used for a number of research and clinical sequencing applications. **(B)** This knowledge can fuel epidemiological studies and allow scientists to characterize major SARS-CoV-2 strains. The green, orange, and pink icons represent contagious individuals who contribute to the transmission of a virus within a population. **(C)** The targeted sequencing enables the detection of low frequency quasispecies that are created through de novo mutations within an individual. The mutation profile from pangenome analysis allow us to examine whether these mutations create a unique genetic profile that can be traced to and across individuals.

RNA-dependent RNA polymerases (RdRp) are the major source of mutations in coronaviruses. Although RNA polymerases are essential for viral replication, this class of polymerases has high error rates, leading to the accumulation of mutations in the viral genome over time [7]. For coronaviruses, estimates of RNA polymerase mutation rates range from 10^−4^ to 10^−6^ mutations per a given nucleotide [8]. Moreover, coronavirus genomes undergo both homologous and nonhomologous recombination which provides additional genetic variation for fueling viral evolution [9]. Although SARS-CoV-2 mutation rates are lower than viruses such as human immunodeficiency virus or influenza [10, 11], the frequency of replication-based mutations is high enough for genetic fingerprinting analysis and its related applications.

Currently, the most common diagnostic methods detect viral RNA, viral antigens, or antibodies produced by the host during an immune response. These molecular detection assays do not provide information about specific viral mutations. Thus, next generation sequencing **(NGS)** is required to identify viral mutations. NGS provides comparable detection sensitivity to the gold standard PCR assays while simultaneously providing viral genome sequence. In addition, sequencing studies of SARS-CoV-2 have generated complete viral genome assemblies, where the entire end-to-end sequence of a viral isolate is reconstructed [12].

With the growing number of sequenced SARS-CoV-2 genomes, researchers have identified numerous mutations indicative of different phylogenetic lineages, otherwise known as clades, and related to specific strains transmitted in the population [13]. Genetic information from these public datasets can be leveraged to trace the spread of infection across different populations. Specifically, one can compare genetic fingerprints using mutation profiles from different viral data sets to map the spread of infections [3], identify virulence factors and study features of viral evolution in populations. Prospectively, the study of these mutations provides a foundation for designing future vaccines and developing drugs for antiviral targets [14].

To date, there are tens of thousands of samples and matched genome assemblies for SARS-CoV-2. With this data, it becomes feasible to conduct ‘pangenome’ studies that simultaneously analyze all of the available viral genome assemblies. The analysis of the SARS-CoV-2 pangenome is useful for determining the common sequence features of all viruses as well as the high frequency mutations that distinguish the viral species most prevalent in an infected population. However, analyzing thousands of viral genome assemblies is time consuming. Among the challenges, there is a lack of common sequence criteria for defining conserved versus hypervariable genomic sequences prone to mutation. Distinguishing between these features is important for understanding the viral evolution, spread and designing future molecular assays that are specific to SARS-CoV-2.

This study had two objectives relevant for viral genetic fingerprinting **(Figure 1)**. The first goal was developing a rapid, efficient method for identifying conserved sequences and mutation profiles across thousands of different viral SARS-CoV-2 genomes in parallel. The second goal was focused on leveraging our results from pangenome analysis to develop a targeted deep sequencing assay. We used it to identify genetic fingerprints characteristic of viral quasispecies from clinical samples and compared these results to the pangenome.

Most SARS-CoV-2 genomic studies have relied on conventional sequence alignment, which is slow and inefficient when applied on a scale of thousands of genome sequences. To overcome this issue and facilitate viral pangenome studies, we developed a rapid computational approach to identify conserved sequences across any number of SARS-CoV-2 genome sequences. This method does not use sequence alignment. Rather, we employ k-mers, short sequences with tens of bases, for annotating and querying the viral genome sequences and genetic variation.

This feature has computational advantages for streamlining the analysis of large number of genome sequences and comparing the features of assemblies. We applied our k-mer approach to find the highly conserved sequences across thousands of SARS-CoV-2 genome assemblies and other viral genome sequence data sets. This process provided the rapid identification of conserved regions of the genome and enabled us to index mutations across thousands of viral genomes.

Based on our identification of conserved regions of the SARS-CoV-2 pangenome, we developed a sequencing assay to identify novel mutations. Rather than covering the entire viral genome, we used highly conserved sequences as primer sites that flank highly variable genome regions. The assay provides very deep sequencing coverage on the average of thousands of reads per a given base. We analyzed a series of contrived samples, artificial viral admixtures and clinical samples. These results from our pilot study showed that one can achieve both high sensitivity and specificity for detecting SARS-CoV-2 with deep targeted sequencing. We detected mutations in the SARS-CoV-2 genome at varying allelic fractions representative of emerging quasispecies. Importantly, when comparing our results with viral mutations from public data sets of viral genomes, our analysis revealed private mutations representing quasispecies unique to a single infected individual. In addition, our multiplexed targeted sequencing assay was significantly easier to implement than whole viral genome sequencing. This multiplexed targeted next generation sequencing assay demonstrated potential for massive scalability required for population studies.

## RESULTS

### K-mer analytics across a SARS-CoV-2 pangenome

To identify conserved regions across thousands of SARS-CoV-2 genome assemblies, we developed a computational workflow that analyzes k-mer sequences **(Figure 1A)**. As previously discussed, k-mers are short sequences of length ‘k’ nucleotides from a given genomic reference, assembly or sequence read data set. This analysis method avoids using multiple sequence alignment and obviates the need for a coordinate system based on one SARS-CoV-2 genome. Conventional alignment techniques are heavily dependent on the given reference genome, and perform comparison between multiple genome sequences across all viral assemblies in a pair-wise fashion. This becomes a very time-consuming process. Furthermore, the complexity of analysis is exacerbated by rapid increases in the number of available viral genome sequences.

As a solution, we developed our approach using 3,968 SARS-CoV-2 genome assemblies, each representing a different viral sample **(Additional file 1; Supplementary Table 1; Additional file 2)**. The sequence assemblies were obtained from GISAID [15]. We built a series of k-mer indices across the entire viral data set. For any given genome, an individual k-mer is derived from single base increments along the length of the viral genome **(Methods)**. Thus, in a genome with a length of 30 kb, the approximate size of the SARS-CoV-2 genome, there are ∼30,000 k-mers. If all viral genomes had the same sequence, this number would not change.

The length of the k-mer sequences was another important variable that we considered. We evaluated k-mers ranging from 21 to 29 bases, the typical length of primers for molecular assays. We chose odd lengths to prevent issues associated with searching for reverse complement sequences. Many of viral k-mer sequences could be identified in the human genome within relatively short edit distances when the length was set at 21 or 23 bases **(Additional file 1; Supplementary Table 2)**. While increasing the k-mer length beyond 25 nucleotides leads to some improvement in terms of specificity for differentiating SARS-CoV-2 sequences from other viral, bacterial and human genome sequences, it reduces the number of conserved and unique k-mers. Balancing these factors, we chose a k-mer length of 25 bases (“25-mer”).

We generated an index of 25-mers from all viral assemblies by associating each 25-mer with the following metadata: 1) IDs of all viral assemblies containing the 25-mer, 2) start position coordinates within each viral assembly containing the 25-mer, and 3) the frequency of each 25- mer within each viral assembly. We identified a total of 94,402 different k-mers from our viral assembly data set. This high number of different k-mers directly reflects viral genome assemblies with mutations. Namely, a variation in sequence such as a mutation generates novel k-mers, thus leading to an increase in the total k-mer number. Without any mutations, we would expect the counts of k-mers around 30,000, which is size of reference SARS-CoV-2 genomes. At this baseline state without additional filtering, the k-mer index corresponded to a matrix M with 94,402 rows (one for each 25-mer), 3,968 columns (one for each SARS-CoV-2 genome), and with elements M[i,j] being the number of times the i-th 25-mer appears in the j-th genome. The k-mer index also contains the locations of each k-mer in each genome, expressed in the local coordinate system per a given genome as metadata. As we demonstrate later, this index enables to generate annotations and compare mutations among different viral genomes very efficiently.

We identified all of the 25-mers that were present only once within an individual viral assembly **(Methods)**. In other words, our definition of a unique k-mer is one where the sequence is found only once in that individual SARS-CoV-2 genome **(Table 1)**. However, the same unique k-mer can also be found in other SARS-CoV-2 assemblies so long as its uniqueness is maintained per a given viral genome **(Figure 1B)**. This simple definition provided us with a way of measuring conservation across different viruses and employing a conservation score. We identified 1,977 k-mers which had the property of being unique amongst all of the k-mers for a given viral genome and where the same 25-mer sequence demonstrated this feature across all of the genomes in our data set (k-mer conservation at 100%). We referred to these conserved sequences as ‘anchor k-mers’ given their properties of both uniqueness and conservation across the pangenome. Additional details about the derivation and properties of the anchor k-mers are described in the **Methods**.

**Table 1:**
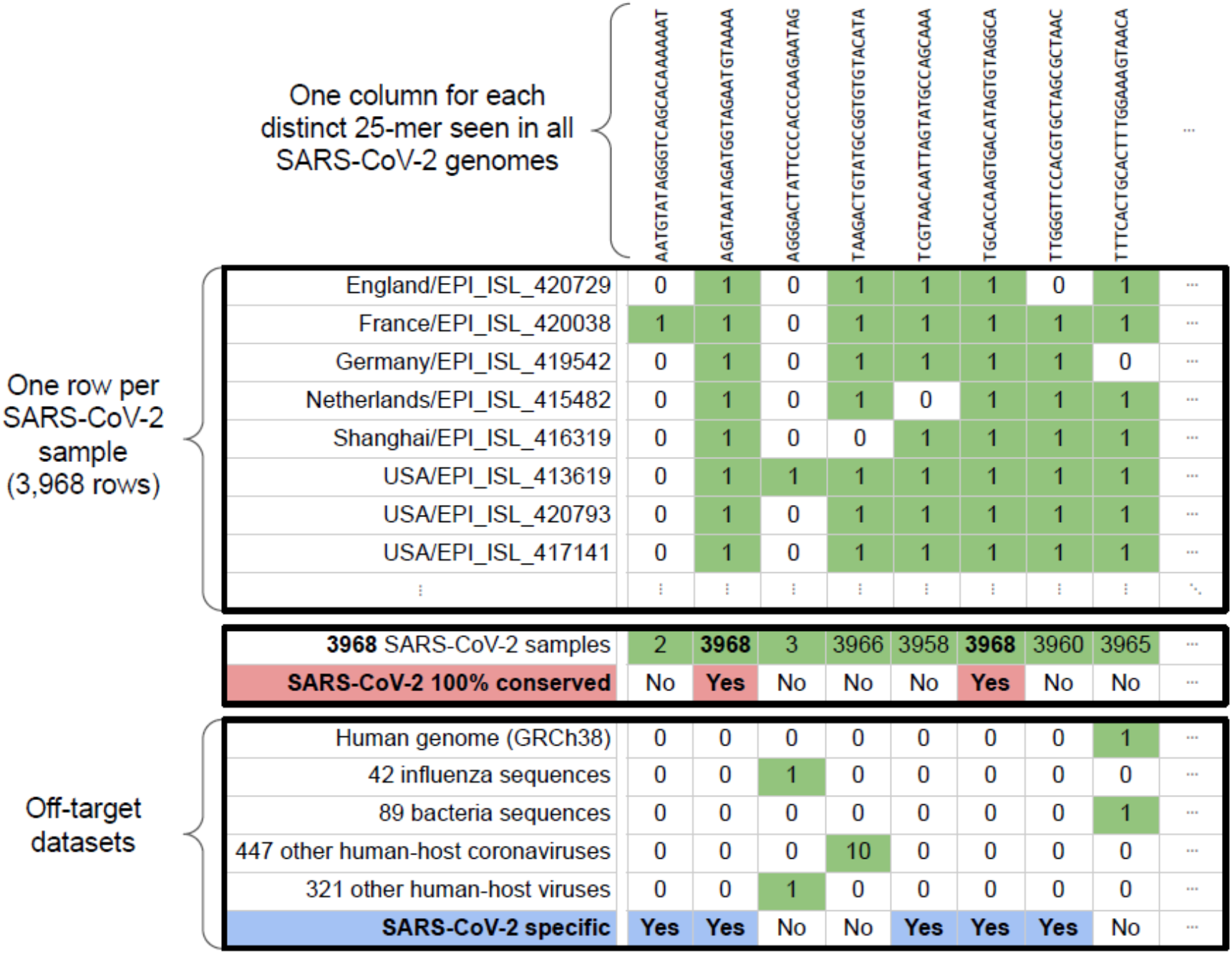
Identification of unique k-mers from viral assemblies.

Our approach was conducted independent of a coordinate system for any given genome and therefore was independent of the choice of a specific SARS-CoV-2 reference. If we conducted this study with an alignment method, comparisons of viral genome assemblies would have scaled exponentially as the number of samples inevitably increased. On the other hand, our method of characterizing the viral pangenome scales linearly and therefore is computationally efficient. Our analysis took a total of five seconds using 32 cores on a server to index approximately 4,000 viral genome assemblies using the k-mer approach.

### Conservation and mutation landscapes of SARS-CoV-2 resolved with k-mer indexing

We grouped the 1,977 anchor 25-mers based on overlapping sequence. As result, we identified 166 highly conserved regions across the 3,968 viral assemblies **(Figure 2)**. The number of overlapping anchor 25-mers in these conserved regions ranges from 1 to 98 with median of 8. The total size of conserved sequences was 5.947kb, which is 19.8% of the 29.9kb reference genome. To display the extent of conserved sequences, we plotted the position of these sequences across the length of a reference viral genome NC_045512.2 for gene and protein annotations **(Figure 2A)**. The gene with the highest number of conserved 25-mers was *orf1ab* **(Figure 2B)**, which is also the largest viral gene. We considered the gene length in the context of the entire genome and normalized the representation as a fraction of the total sequence. By correcting for sequence length, we found that *orf7a* and *orf7b* were the most highly conserved at 24.96% and 30.3%, respectively.

**Figure 2.**
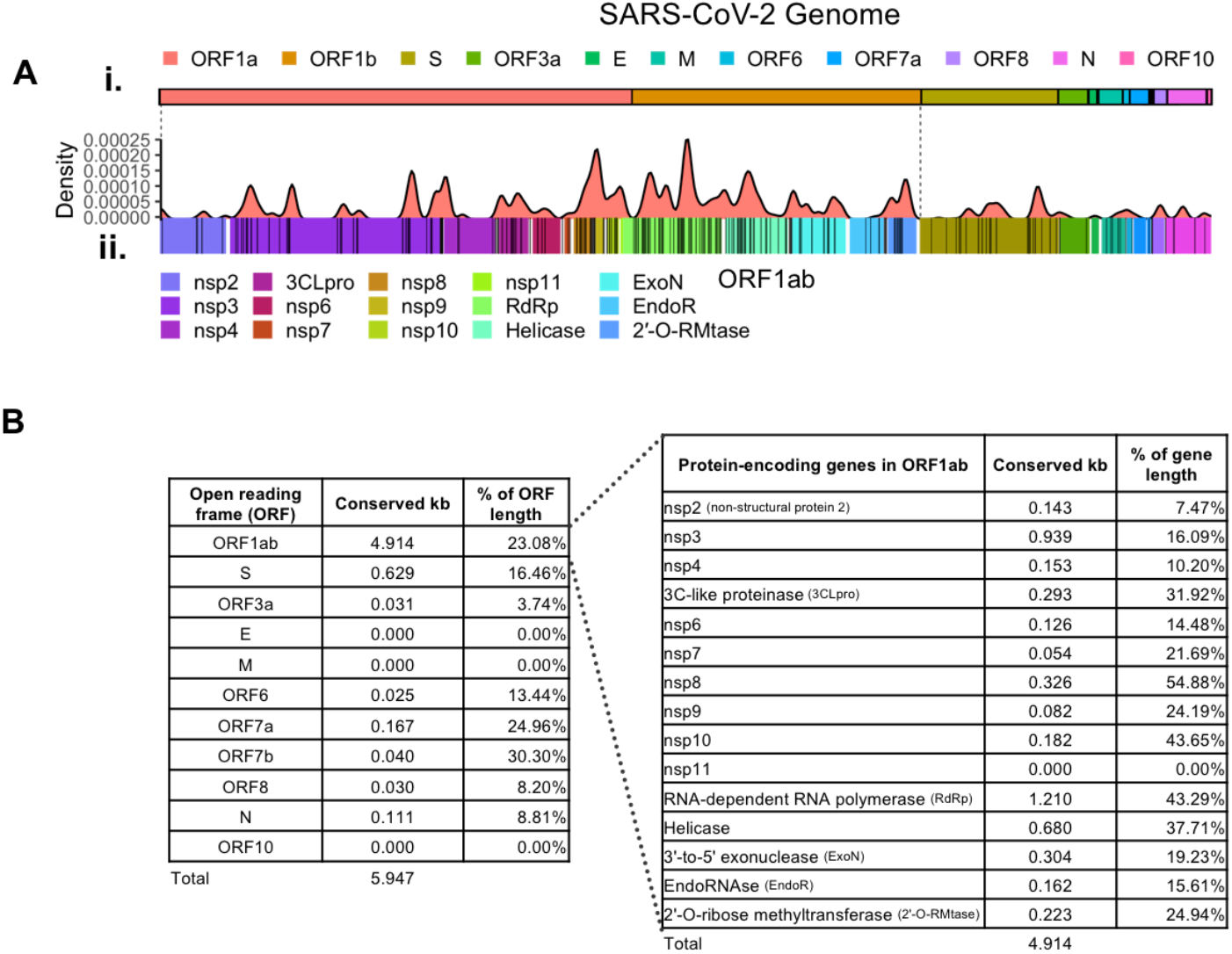
SARS-CoV-2 landscape of conserved regional sequences. A total of 1,977 conserved 25-mers (“anchor 25-mers”) were identified from the 3,968 SARS-CoV-2 genome sequences included in the k-mer analysis. (**A)** Distribution of anchor 25-mers across the (i) SARS-CoV-2 genome. Individual anchor 25-mers are shown as (ii) black lines with their overall density. Conserved regions consist of either discrete or overlapping anchor 25-mers and vary in total length (black rug plot). The density of conservation refers to the total number of base pairs comprising anchor 25-mers within each consecutive 100 bp window across the genome (red kernel density plot). **(B)** The total number of conserved base pairs within each given region of the genome varies across ORFs and genes.

The *orf1ab* sequence encodes multiple proteins **(Figure 2B)**. Interestingly, three protein-coding regions within *orf1ab* had high percentage of conserved sequences: 54.88% for nsp8; 43.65% for nsp10; 43.29% for RdRp. In contrast, the *E* gene did not overlap with any conserved regions. This result suggests that the *E* gene is more likely to subject to significant mutations during the course of viral evolution.

From our initial pangenome dataset of 3,968 SARS-CoV-2 genomes, we identified 2,346 mutations occurring in at least one SARS-CoV-2 genome assembly **(Additional file 3)**. Out of these variants, 1,005 (42.8%) were non-synonymous mutations, and 1,577 mutations (63.9%) were unique to a single isolate. Thirty-eight mutations were found in more than 1% of genomes, while 15 mutations were found in more than 5% of genomes. The most common mutations found in more than 40% of genomes were as follows: 1) 14408 C>T; RdRp P323L (45.7%), 2) 3037 C>T; nsp3 F106 (45.3%), 3) 23403 A>G; S D614G, and 4) 241 C>T; 5’ UTR (45.2%).

We further conducted an expanded pangenome analysis by examining an additional set of 75,681 complete and high-coverage viral genomes from GISAID (downloaded date: 09/23/2020). These mutations were distinguished by patterns of unique 25-mer sequences **(Supplementary Figure 1)** and were incorporated into our k-mer mutation index. We identified 20,671 mutations from at least one viral genome assembly; 1) 15,914 of these mutations (21%) were found in less than 0.001% of genomes and 2) 8,071 (10.6%) were unique to a single sample **(Figure 3A, Additional file 3)**. On average, each viral genome contains 8.61 mutations with a standard deviation of 3.45.

**Figure 3.**
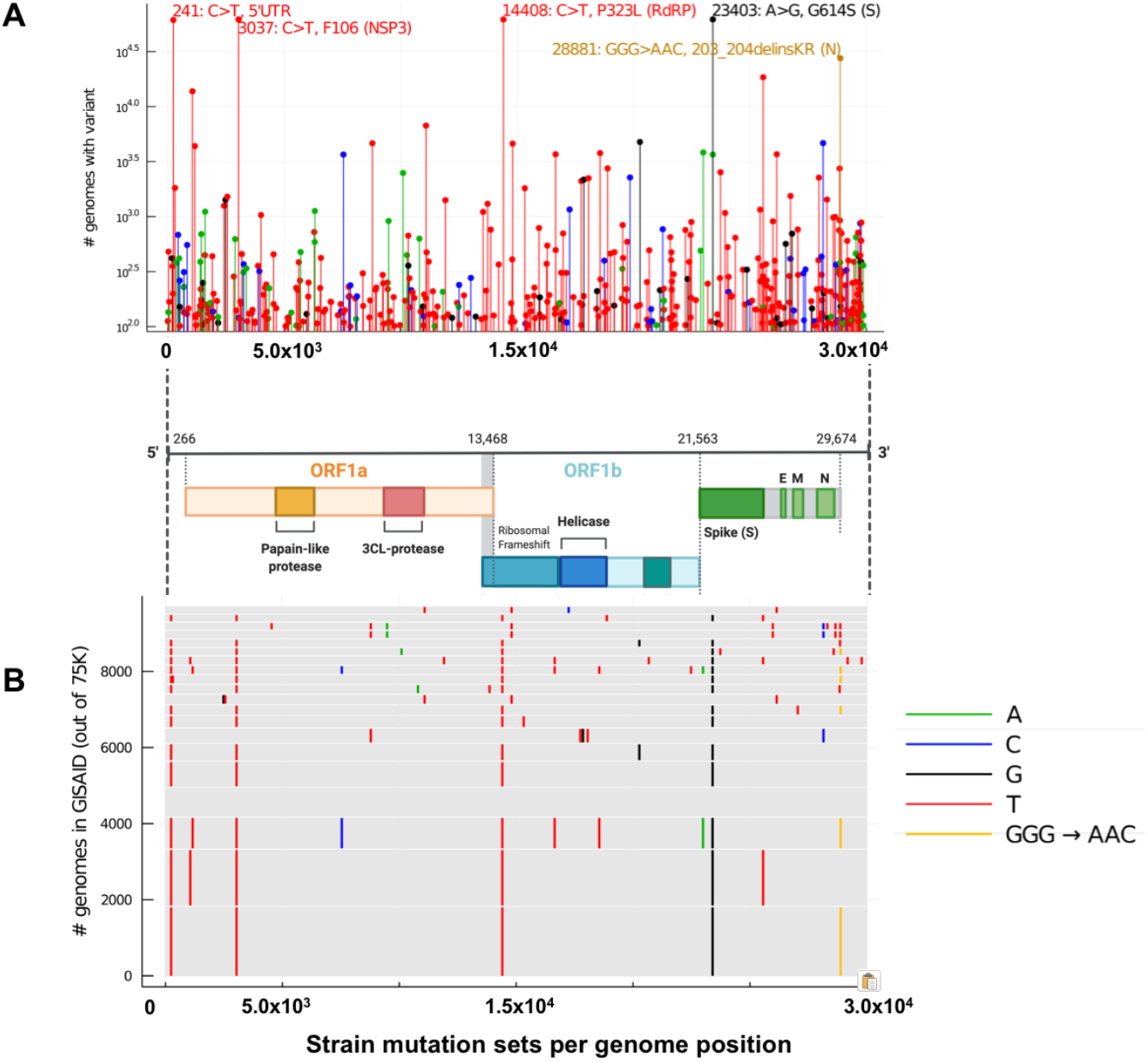
Individual and joint frequencies of mutations in 75,681 SARS-CoV-2 genomes. **(A)** Mutations occurring in at least 100 of the 75,681 SARS-CoV-2 genomes, with top 5 most frequent mutations annotated. Colors indicate the variant allele (see legend). **(B)** Top 20 most frequent co-occurring sets of mutations (“fingerprints”) among 75,681 GISAID genomes. Each gray rectangle corresponds to one fingerprint, with vertical line segments located at the positions of the variants in that fingerprint, and with line segment colors indicating the variant alleles (see legend). The height of each rectangle is equal to the number of GISAID genomes with the associated fingerprint, so the single most frequent fingerprint is the bottommost rectangle. The 4th most frequent fingerprint (4th rectangle from the bottom) has no variants relative to the SARS-CoV-2 reference genome (NC_045512).

From this expanded pangenome analysis, we identified mutation profiles that were highly specific viral identifiers, or genetic “fingerprints,” based on their relatively low frequency among these samples. Among the notable characteristics, 12,552 (60.7%) mutations were nonsynonymous, 7,234 (35%) were synonymous and 885 (4.3%) mutations occurred within non-coding regions, such as the 5’ UTR and the 3’ UTR. Fifty-seven mutations were found in greater than 1% of assemblies while 14 variants were found in greater than 5% of assemblies. We found that *RdRp* had the largest number of mutations that occurred more than 1% of assemblies (7 out of 57 mutations). The most common mutations we observed were as follows: 62,018 (81.95%) for 3037C >T (F106) in the nsp3 protein; 61,993 (81.91%) for the 23404A>G (D614G) in the *S* gene; 61,920 (81.82%) for 14408C>T (P323L) in the RdRp protein **(Figure 3A)**.

Using our k-mer mutation index, we identified the twenty most common combination profiles of mutations, or “fingerprints,” among the 75,681 genome assemblies that we examined **(Figure 3B)**. As shown in **Figure 3B**, the top 20 SARS-CoV-2 genetic fingerprints represented the mutation profile of approximately 11.6% of viral assemblies. The remaining 88.4% had more discrete sets of mutations, suggesting that these signatures occurred less frequently across the viral genomes included in our analysis. Notably, the fourth most frequent fingerprint (occurring in approximately 1% of assemblies included in our analysis) had no mutations relative to the SARS-CoV-2 reference genome (**Figure 3B**). The overall majority of viruses included in our analyses had at least one viral genetic signature with the vast majority occurring at low frequencies.

### Quantitative measurement of SARS-CoV-2 with deep targeted sequencing

Based on our pangenome results, we designed a targeted sequencing assay using the highly conserved sequences from anchor 25-mers. Our goal was to develop a robust amplification assay covering a wide portion of the viral genome but not necessarily the entire viral genome. This latter point was important, seeing that we intended to generate sequencing coverage in the thousands at a reasonable sequencing cost and with a minimal number of amplicons.

As part of the amplicon design, we considered the following parameters for primer sequences: (1) specific only to SARS-CoV-2; (2) present in highly conserved regions per our pangenome analysis; (3) flanking non-conserved regions; (4) having DNA properties such as GC content that facilitated multiplex PCR; (5) minimizing the number of amplicons to reduce multiplexing artifacts. Based on these combined parameters, we anticipated that our primer sets had a very high probability of amplifying any SARS-CoV-2 genomes regardless of the location of any mutations. As illustrated in **Figure 2**, we found a total of 1,977 anchor 25-mers, all being highly conserved across nearly 4,000 viral genomes. Interestingly, when we examined the ARTIC primers which are commonly used for viral genome sequencing [16], only 57% of the primers appeared in at least 99% of the 75,681k GISAID dataset.

We followed the Food and Drug Administration’s **(FDA)** Emergency Use Authorization guidelines for designing SARS-CoV-2 detection assays. The FDA’s criteria include testing for cross-reactivity against other viral, bacterial, and human genomes **(Additional file 1; Supplementary Table 3)**. To reduce the chance of off-target amplification, we eliminated any candidate primer sequences up to an edit distance of four present in other human coronavirus, human viruses and bacteria. Likewise, we used the same criteria for sequences that were found in the human genome and excluded any candidate primer sequences within an edit distance of two **(Methods)**.

This multiplexed assay had six amplicons targeting the more variable regions of the SARS-CoV- 2 genome, for a total coverage of approximately 39.9% of the viral genome **(Figure 4A; Additional file 1; Supplementary Table 4)**. Amplicon sizes ranged from 1kb up to 2.67kb. In addition, we amplified a region in human gene *RPP30* as a positive control against technical problems that may occur during RNA extraction. Polymorphisms detected in this gene also serves as a control against potential sample swaps or contamination.

**Figure 4.**
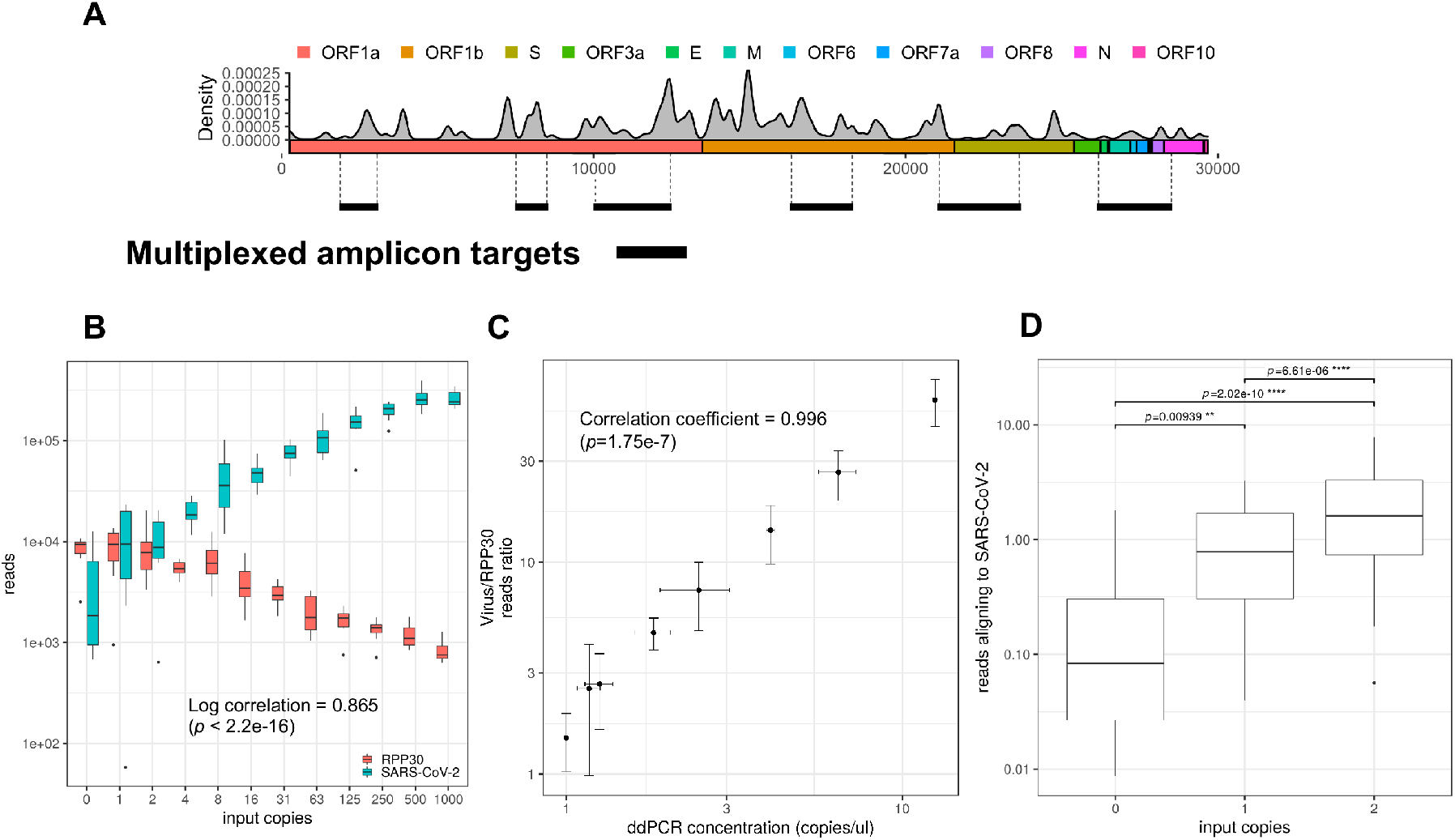
Deep targeted sequencing of SARS-CoV-2 by multiplex amplicon generation. **(A)** Amplicon targets. Six amplicons (dark line) ranging from 1kb to 2.6kb were used to target approximately 40% of the SARS-CoV-2 genome. The density of anchor 25- mers indicating high sequence homology across GISAID isolates are also shown. **(B)** Testing dynamic range of SARS-CoV-2 sequencing. Next-generation sequencing libraries were created from the SARS-CoV-2 amplicons as well as targeting the *RPP30* control gene. Libraries consisted of serial dilutions (N=8 replicates) of purified SARS-CoV-2 genomic RNA into total nucleic acid derived from a COVID-19 negative saliva sample. Reads aligning to either *RPP30* or SARS-CoV-2 are shown, alongside with the log-log correlation to the number of input copies of virus in each sample. **(C)** Correlation with digital PCR. At each dilution used for sequencing, a portion of cDNA was used for digital PCR (N=3 replicates).The corresponding concentration derived from digital PCR is shown alongside reads aligning to virus versus the *RPP30* control (correlation coefficient=0.996, p<1.75e-7). **(D)** Limit of detection testing. A larger number of replicates (N=32) was performed at 0, 1, and 2 input viral copies to assess the limit of detection of the assay. Shown are the reads aligning to both SARS-CoV-2 and the *RPP30* control gene. Significance values (FDR-corrected) which test against the negative control (0 copies) are shown.

First, we used randomly primed reverse transcription of viral and human RNA. This step was followed by a multiplexed PCR amplification. Afterwards, to generate Illumina-compatible sequencing libraries we used a two-step transposase approach where the first step incorporates a unique DNA barcode among 96 wells and the second pooled reaction adds a plate-specific barcode **(Methods)**. For the first step, the amount of tagmentation conducted by transposase is limiting for any given reaction volume. Thus, sequencing libraries are automatically normalized prior to loading onto a sequencing instrument, reducing the total hands-on operational time. With these new features, our experimental approach was highly scalable, enabling the generation of hundreds of normalized sequencing libraries in a single day without the need for robotic automation.

We tested this sequencing assay on cDNA generated from a serial dilution of twelve different concentrations of commercially available SARS-CoV-2 genomic RNA **(Figure 4)**. The serially-diluted SARS-CoV-2 RNA was spiked into total nucleic acid extracted from human saliva. The human RNA sample was SARS-CoV-2 negative as per qPCR. The range of SARS-CoV-2 concentrations spanned three orders of magnitude from 1,000 total copies to 1 total copy per a given sample **(Additional file 1; Supplementary Table 5)**. We generated and sequenced eight technical replicates per concentration from the cDNA samples. Following Illumina sequencing, the data was processed and aligned to both the human genome (GRCh38) and the SARS-CoV- 2 reference NC_045512.2. Overall, the coverage in viral regions was over 1,000X with high viral inputs with over 95% of target regions sequenced, but declined as viral inputs decreased towards single viral copies **(Additional file 4)**. There was a high correlation between sequence reads aligning to the SARS-CoV-2 genome compared to the amount of viral RNA **(Figure 4B)** with a log-log correlation coefficient of 0.865 (*p* < 2.2e-16). In parallel, as the viral input decreased, the proportional number of *RRP30* reads increased. Here, we found that deep targeted sequencing enabled sensitive detection of SARS-CoV-2.

To validate the quantitative performance of the sequencing assay, we performed digital PCR **(dPCR)** on the same batch of cDNAs used in the SARS-CoV-2 dilution series **(Figure 4C)**. Digital PCR provides absolute quantitative measurement of RNA templates at near single molecule resolution. We used a set of PCR primers for the N1 region and a TaqMan-based oligonucleotide probe to detect the presence of template in each droplet **(Methods)**. We observed a high concordance between digital PCR and sequencing (Pearson correlation coefficient = 0.996; *p*=1.75e-7; log-transformed values), supporting equivalent sensitivity between the two methods **(Figure 4D)**. Overall, we demonstrated sequencing-based viral detection at a resolution as low as a single copy per reaction.

We further validated the lower detection threshold of sensitivity of our sequencing assay. For this experiment, we used the cDNA samples with absolute quantitation from dPCR. We generated 32 amplicon replicates from cDNA samples containing two, one, and zero SARS-CoV-2 genomic copies **(Figure 4D)**. We detected observed statistically significant differences between one and zero input copies (p=0.0094; one-sided FDR-corrected t-test), suggesting that this sequencing assay may have more sensitive detection compared to other molecular diagnostic assays for SARS-CoV-2 detection [17].

### Detection of viral mutation allelic fractions

We validated our sequencing assay for detecting mutations at low read frequency and resolving genetic fingerprints. We generated admixtures of SARS-CoV-2 RNA from two strains originating from separate continents (USA-WA1/2020 and Hong Kong/VM20001061/2020) to test the capability of the k-mer-based mutation index to distinguish mutations among different viral strains. Within the targeted regions that we sequenced, the Hong Kong strain had a total of six exclusive mutations and the Washington state strain had one exclusive mutation **(Additional file 1; Supplemental Table 6)**. We prepared viral RNA admixtures with different relative fractions of each strain for a total input of 1,000 copies, which were spiked into human nucleic acid. The admixture ranged from a high of 99% to a low value of 1% for a given viral strain component. Eight technical replicates of each admixture were prepared and sequenced in parallel. The average sequencing coverage for all the replicates was in the thousands **(Additional file 4)**. Using the published assembly sequences of each strain and their associated mutations, we determined the read coverage of each mutation base **(Additional file 1; Supplemental Table 7)**. We plotted with the empirical value versus the expected allelic fraction **(Figure 5)**. In the case of the mutations with the Hong Kong strain, the correlation coefficient was 0.983. For the Washington strain, the correlation coefficient was 0.998. Our results showed that the measured mutation allelic fraction per our sequencing data highly correlated with the theoretical admixture fraction. Moreover, mutations expected to be present at a 1% allelic fraction were detected across all replicates. This result indicates that sensitive detection of low allelic fraction mutations was feasible.

**Figure 5.**
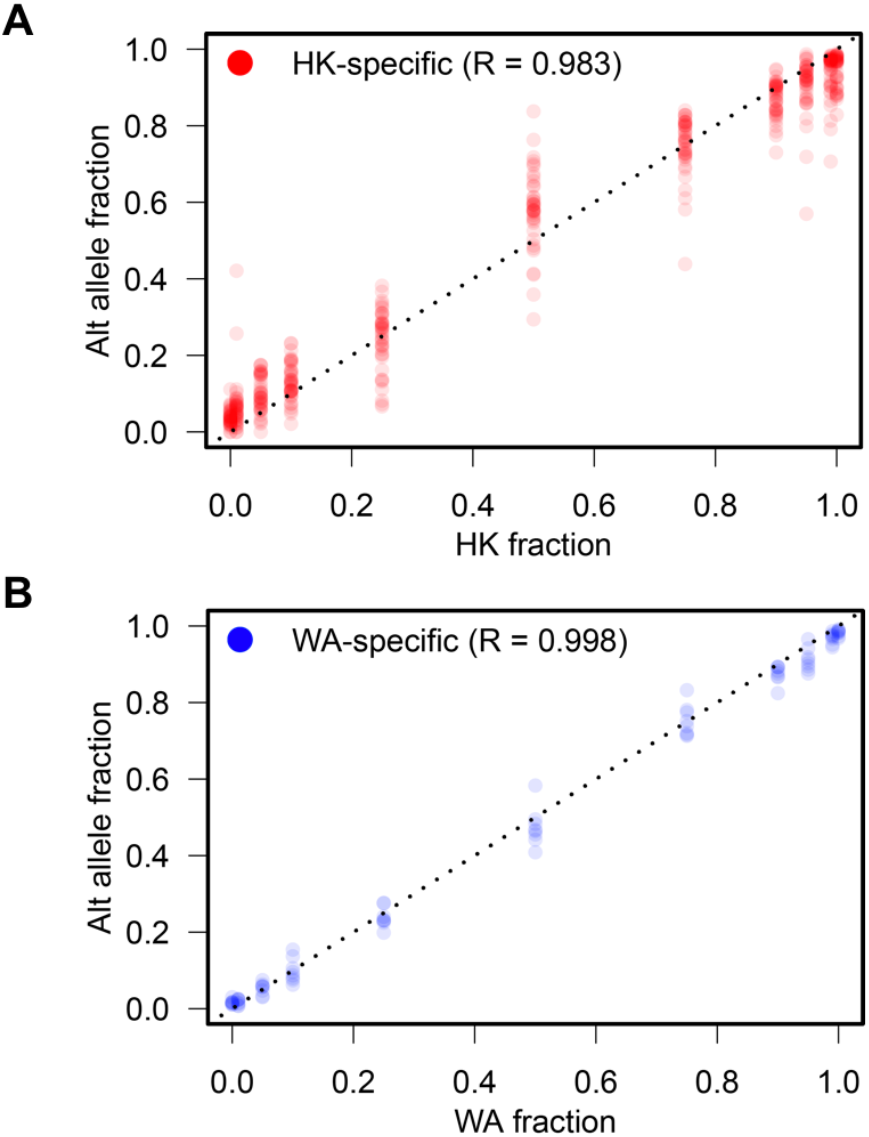
Fraction of alternative alleles specific to viral strains in admixtures. The allelic fraction of exclusive mutations, measured from all the eight replications, were compared against the ratio of viral strains, separately for the Hong Kong **(A)** and the Washington **(B)** strains. The admixtures contain 0%, 1%, 5%, 10%, 25%, 50%, 75%, 90%, 95%, 99%, or 100% of either strain (the x-axes). The allelic fraction of exclusive mutations are plotted on the y-axes. Areas of the plot with many overlapping points appear as dark clusters, and the dotted diagonal line indicates 1:1 concordance. For both the strains, the Pearson’s correlations (R) are indicated.

### Viral quasispecies analysis of a SARS-CoV-2 patient cohort

We examined a set of SARS-CoV-2 clinical samples that include both positive and negative samples previously evaluated with a RT-qPCR assay (FDA EUA200036) approved by the US Food and Drug Administration **(FDA)**. We sequenced a total of 100 extracted RNA clinical specimens from nasopharyngeal swabs obtained from patients tested at the Stanford Clinical Virology lab. The specimens consisted of 30 clinically diagnosed positives and 70 clinically-diagnosed negatives **(Additional file 8; Supplemental Table 8)**. A total of three cDNA technical replicates were made from each of the 30 positive RNA samples for immediate downstream use in PCR amplicon generation and sequencing library preparation.

Overall, each sample yielded high quality reads, of which Q30 scores were over 90%. In addition, 99% of the reads aligned to either the human or SARS-CoV-2 genome **(Additional file 4)**. We observed that some samples had substantially lower sequencing yield. Upon further investigation, these samples had low numbers of reads in both viral and human targets across replicates, indicating low viral RNA yield from nasopharyngeal swabbing or sample extraction. Viral sequencing coverage varied from less than 100X to greater than 10,000X, indicating large-scale variation in viral load across patients.

While developing the assay, we observed that the proportional number of reads aligning to either SARS-CoV-2 or *RPP30* was dependent on the number of viral copies loaded. Therefore, we utilized the virus-to-*RPP30* read ratio as a normalization metric to assess performance and detection **(Figure 6)**. We found that this metric had a high correlation to the Ct values from qPCR diagnostic test (Pearson correlation coefficient of -0.90, *p*=2.4e-11) **(Figure 6; inset)**. Previously found to be SARS-CoV-2 negative by qPCR, these 70 clinical samples provided us with a calibration threshold for determining a positive case. We determined the average read ratio and standard deviation to be 0.00494 and 0.00163, respectively. We set our threshold at three standard deviations (0.00488) above the average to detect the presence of SARS-CoV-2 reads. Based on this definition, the sequencing results were positive for 30 out of 30 samples previously diagnosed by qPCR. All negative samples had a read ratio of less than 2%, well below the read ratio of the positive samples. A small proportion of viral reads were noted in the negative controls and samples. We suspect these viral reads were an artifact in which sample indexes are swapped among multiplexed libraries [18].

**Figure 6.**
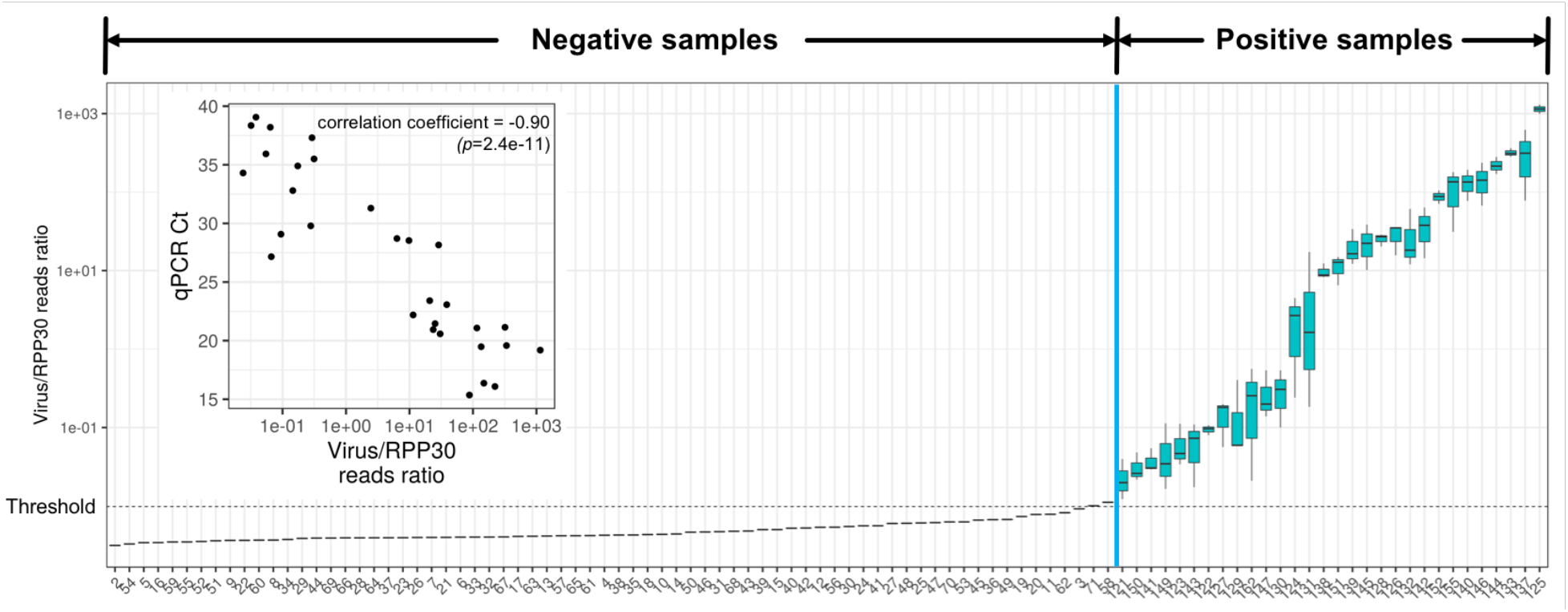
Targeted sequencing of SARS-CoV-2 of clinical samples. Targeted sequencing was performed on a total of 100 clinical samples (30 positive and 70 negative) derived from nasopharyngeal swabs. The ratio of reads aligning to viral and *RPP30* sequences is displayed, with samples shown in increasing order of the ratio of reads aligning to SARS-CoV-2 to *RPP30*. The positive samples (blue; N=3) were found to be higher than the reads ratio of negative samples by setting a threshold of greater than three times greater than the standard deviation away from the mean of the negative samples (dashed line). Inset: Correlation of viral reads ratio to qPCR CT values as derived from clinical tests (correlation coefficient=-0.90, p=2.4e-11).

After sequence data processing, we identified mutations from the 30 COVID-19 positive clinical samples and a control SARS-CoV-2 isolate **(Methods)**. We identified a total of 37 mutations across 26 positions from 16 out of the 30 clinical samples **(Table 2)**. The remaining 14 samples showed no variant differences from the reference assembly NC_045512.2. We observed a broad range of mutation allelic fractions, ranging from less than 10%, indicative of subclonal mutations to strain-specific mutations, which were greater than 90% for a given viral sample **(Figure 7A)**. We observed 25 positions with substitutions (16 missense and 9 silent) and 1 position with a deletion **(Figure 7B)**. Mutations leading to substitutions were noted at 20 of the 26 total positions. These mutations were unique to an individual sample. The remaining six substitutions were found in more than one sample **(Table 2)**. Among our samples, six mutations were also found in with high frequency (greater than 1,000 samples) in different data set such as the WHO SARS-CoV-2 (N=10,022) and the GISAID (N=75,681) collection. Four mutations were observed in the SARS-CoV-2 genomic RNA isolate from Washington which we used as a positive control. Interestingly, two of these four mutations were shared with one patient sample (P145) and two were private to the positive control.

**Table 2:** Observed SARS-CoV-2 mutations in clinical samples.

| **Mutation** | **Gene / Open Reading Frame** | **Translational change** | **Sample frequency (n=30)** | **Samples** | **GISAIID frequency (n=75,681)** |
| --- | --- | --- | --- | --- | --- |
| 2721 C > T | Nsp3 | A1V | 1 | P145 | 7 |
| 3037 C > T | Nsp3 | synonymous | 5 | P133, P152, P128, P140, P125 | 62018 |
| 7482 C > T | Nsp3 | S1588L | 1 | P162 | 10 |
| 8092 C > T | Nsp3 | synonymous | 1 | P145 | 42 |
| 8389 C > T | Nsp3 | synonymous | 1 | P133 | 154 |
| 11074 CT > C | NA | microsatellite | 3 | P155, P128, P142 | NA |
| 16267 C > A | Helicase | Q11K | 1 | ATCC_WA-01 | NA |
| 16375 C > T | Helicase | P47S | 1 | P142 | 16 |
| 16876 A > G | Helicase | T214A | 1 | P132 | 0 |
| 17747 C > T | Helicase | P504L | 1 | P145 | 2104 |
| 17858 A > G | Helicase | Y541C | 1 | P145 | 2167 |
| 18060 C > T | 3' to 5' exonuclease | synonymous | 2 | ATCC_WA-01, P145 | 2233 |
| 18084 C > T | 3' to 5' exonuclease | synonymous | 1 | P126 | 4 |
| 21646 C > T | S | synonymous | 1 | P151 | 257 |
| 22289 G > T | S | A243S | 1 | P152 | 15 |
| 23403 A > G | S | D614G | 3 | P152, P125, P133 | 61993 |
| 26542 C > T | M | T7I | 1 | ATCC_WA-01 | 13 |
| 26625 C > T | M | synonymous | 1 | P144 | 86 |
| 26951 G > T | M | synonymous | 1 | P146 | 44 |
| 27131 C > T | M | synonymous | 1 | P137 | 16 |
| 27641 C > T | ORF7a | S83L | 1 | P126 | 31 |
| 27670 G > T | ORF7a | V93F | 1 | P139 | 35 |
| 27874 C > T | ORF7b | T40I | 2 | P144, P146 | 40 |
| 27925 C > T | ORF8 | T11I | 1 | P138 | 22 |
| 27970 C > T | ORF8 | T26I | 1 | P132 | 66 |
| 28144 T > C | ORF8 | L84S | 2 | P145, ATCC_WA-01 | 4655 |

**Figure 7.**
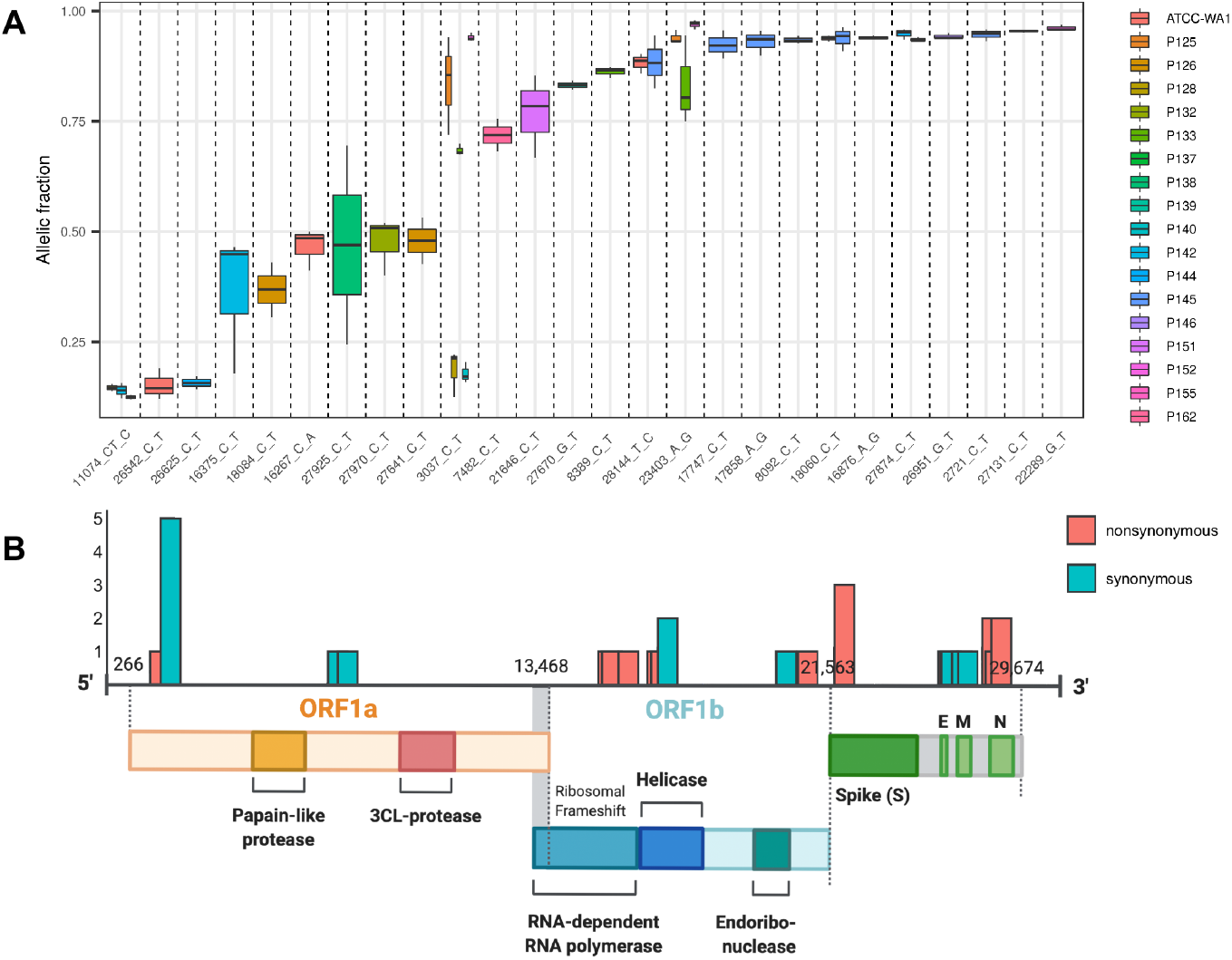
Quantifying viral quasispecies signatures in clinical samples. **(A)** Allelic fraction across mutations. Each boxplot represents the distribution of allele fractions for each viral sample at a particular viral genome position. Some mutations were found in multiple viral samples and are so noted with multiple boxplots. **(B)** Distribution of mutations across the viral genome. The number of mutations found at each position, as well as its class (synonymous vs. nonsynonymous) is shown.

We observed five mutations, each private to a single sample, in the helicase gene from three clinical samples (Table 2). One sample (P145) had two mutations in the helicase gene and the other two (P142 and P132) had only one mutation. The two helicase mutations (17747 C > T and 17858 A > G) found in sample P145 were reported in 2,104 (2.78%) and 2,167 (2.86%), respectively, out of the 75,681 GISAID genomes included in our analysis. The other mutations observed in the helicase gene were present in less than 0.03% of the 75,000 genomes annotated in GISAID genomes.

We identified three nonsynonymous mutations in *orf7* and three nonsynonymous mutations in *orf8* from the clinical specimens. All but one (27874 C > T in *orf7b*) were private to their respective samples **(Table 2)**. Two of the mutations in *orf7* were in *orf7a* (P126 and P139) and one was in *orf7b* (P144 and 146). All three *orf7* mutations were present in less than 0.1% of the SARS-CoV-2 genomes included in our pangenome analysis. In *orf8*, one mutation (28144 T > C) was identified in a clinical sample (P145), present in the positive control strain (ATCC_WA- 01) and observed in 4,655 genomes, or 6.15% per GISAID. The other mutations in *orf8* were seen in frequencies less than 0.1% in the GISAID pangenome analysis **(Table 2; Supplementary Table 9)**. Interestingly, all of the mutations observed in both *orf7* and *orf8* among our clinical sample set are expected to result in significant changes to the amino acid side chain polarity **(Table 2)**. For example, three of the six mutations observed between *orf7* and *orf8* (27874 C > T in samples P144 and P146, 27925 C > T in sample P138, 27970 C > T in P132) lead to an amino acid change from threonine to isoleucine which is a major change in sidechain polarity.

### Comparisons of SARS-CoV-2 genetic fingerprints

We compared our mutation profiles among the 17 patient samples with strain-specific or quasispecies-level mutations representative of subclonal viral populations. Hierarchical clustering of mutations points out some potential group structure **(Figure 8)**. While some individual mutations were shared between two or more samples, no two samples had the same mutation profile. In other words, each patient had their own unique genetic viral fingerprint with no direct match among our set of clinical samples. Notably, we observed private mutations for each sample. Three clinical viral samples had mutations which occurred in less than 0.01% of 75k GISAID viral samples.

**Figure 8.**
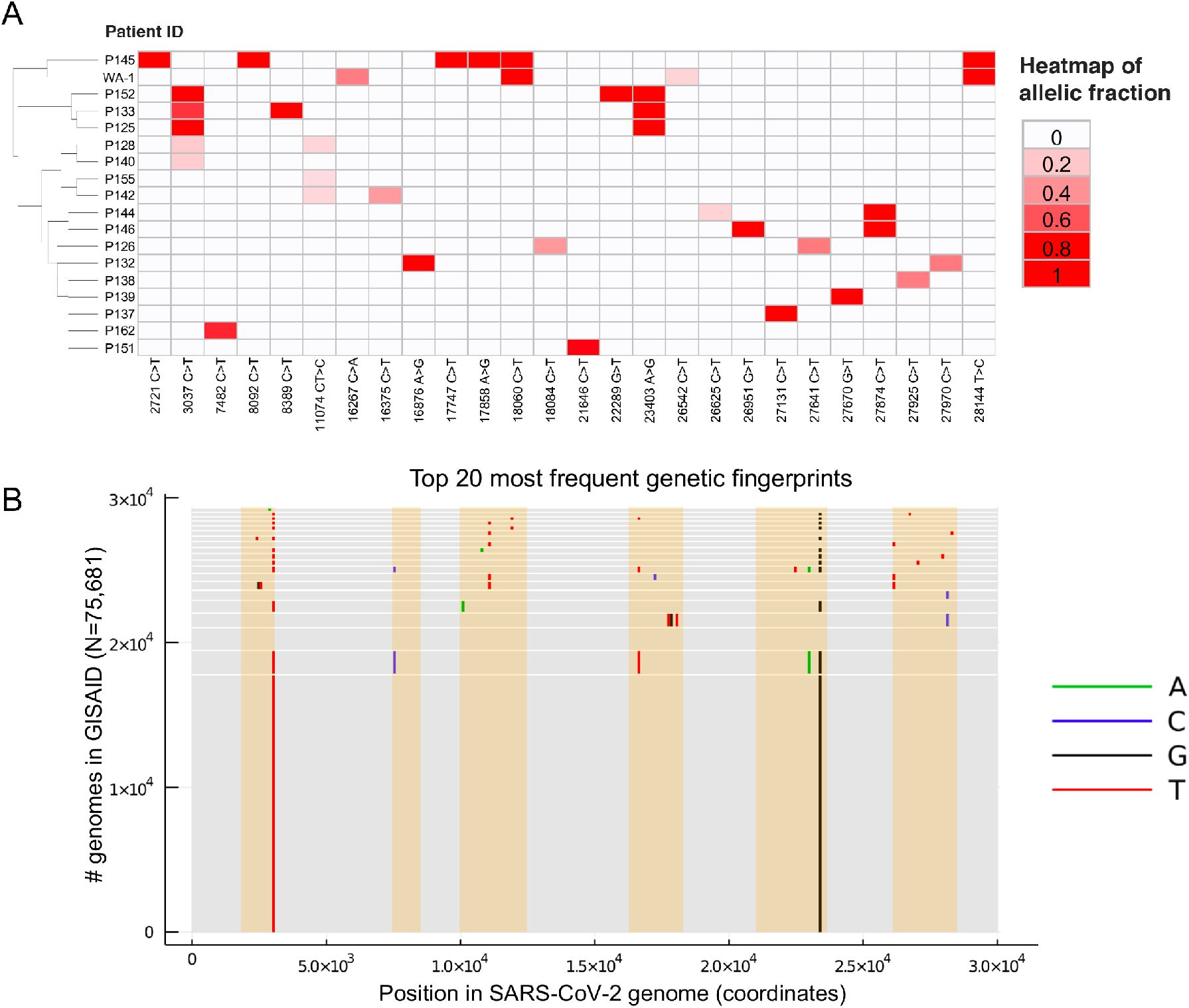
Heat map of allelic fractions in positive clinical samples. **(A)** Rows and columns indicate samples and the mutations, respectively. Result from a hierarchical clustering is shown as dendrogram. To best represent evolutionary relationship among samples, the clustering was based on the genotype itself (i.e. mutation or no mutation) instead of the allelic fraction values. The mutations are sorted by the genomic position. **(B)** Top 20 most frequent co-occurring sets of mutations (“fingerprints”) among 75,681 GISAID genomes, with variants restricted to lie inside the regions of the SARS-CoV-2 genome that we sequenced. Each gray rectangle corresponds to one fingerprint, with vertical line segments located at the positions of the mutation in that fingerprint, and with line segment colors indicating the mutation alleles. The height of each rectangle is equal to the number of GISAID genomes with the associated fingerprint, so the single most frequent fingerprint is the bottommost rectangle. The 3rd most frequent fingerprint (3rd rectangle from the bottom) has no variants relative to the SARS-CoV-2 reference genome within the sequenced regions.

As we described previously, we constructed a k-mer mutation index database in which individual mutations were represented for 75,681 viral samples from GISAID. These mutations had passed quality control criteria to insure a higher likelihood of being accurate **(Methods)**. The k-mer index facilitated rapid pairwise similarity calculations among our samples versus those from the GISAID **(Table 3)**. When making comparisons from our clinical cohort with this expanded set of samples, we used two different criteria between our viral sample set that underwent deep sequencing versus the mutation signatures from GISAID **(Table 3)**. First, we applied strict matching criteria such that a mutation set per a given Stanford clinical sample exactly matched the set of another sample annotated in GISAID. When using the strict matching criteria, there were seven samples that were unique and not found among the 75,681 GISAID samples. In addition, there were four patients (P137, P139, P146, P152) with viral samples which did match those in GISAID but were present at a frequency of less than 0.0001%.

**Table 3:** Viral mutation fingerprint comparisons.

| Patient | Mutations found in patient viral sample |  | GISAID viral genomes (N=75,681) |  |  |
| --- | --- | --- | --- | --- | --- |
|  | Private mutation<sup>1</sup> | Reported mutations | Strict matching # genomes<sup>3</sup> | Relaxed matching # genomes<sup>4</sup> | Ratio relaxed criteria |
| P125 |  | 3037C>T, **23403A>G** | 17851 | 43943 | 5.806345E-01 |
| P126 | 18084C>T | **27641C>T** | 0 | 0 | 0.000000E+00 |
| P128 |  | **3037C>T** | 66 | 61952 | 8.185938E-01 |
| P132 | 16876A>G | **27970C>T** | 0 | 0 | 0.000000E+00 |
| P133 |  | **3037C>T**, 8389C>T, **23403A>G** | 37 | 115 | 1.519536E-03 |
| P137 |  | 27131C>T | 1 | 15 | 1.982003E-04 |
| P138 |  | **27925C>T** | 0 | 22 | 2.906938E-04 |
| P139 |  | **27670G>T** | 1 | 34 | 4.492541E-04 |
| P140 |  | **3037C>T** | 66 | 61952 | 8.185938E-01 |
| P142 |  | **16375C>T** | 0 | 16 | 2.114137E-04 |
| P144 |  | 26625C>T, **27874C>T** | 0 | 0 | 0.000000E+00 |
| P145 | **2721C>T** | 8092C>T, **17747C>T**, **17858A>G**, 18060C>T, **28144T>C** | 0 | 0 | 0.000000E+00 |
| P146 |  | 26951G>T, **27874C>T** | 3 | 0 | 0.000000E+00 |
| P151 |  | 21646C>T | 0 | 257 | 3.395833E-03 |
| P152 |  | 3037C>T, 22289G>T, 23403A>G | 4 | 9 | 1.189202E-04 |
| P162 |  | **7482C>T** | 0 | 10 | 1.321336E-04 |

1. Private mutations refer to those with a frequency less than 0.001%
2. Mutation in bold are non-synonymous
3. Strictly refers to other samples with the exact same number of mutations
4. Relaxed refers to samples that are inclusive for these mutations but may have others

Given that we only had partial genome coverage, we considered a second criterion in assessing the genetic fingerprints observed in patient samples **(Table 3)**. We had initially required that the mutation signatures from the clinical samples have an exact mutation set match. Under our new relaxed matching criteria, we also included GISAID samples with mutations additional to those identified in our sequencing study. Given that the alignments downloaded from GISAID were obtained via whole genome sequencing, this allowed us to include samples with mutations outside of the regions we targeted.

When we considered the relaxed criteria, there were four samples which had no matches with GISAID (P126, P132, P144, P145). Nine samples (P133, P137, P138, P139, P142, P146, P151, P152) had viral GISAID matches, but when considering them as a subtotal, these sets of mutations occurred at a frequency of less than 0.001% among GISAID genomes included in our analysis. There were three patients (P125, P128, P140) in which, when using the relaxed matching criteria, the frequency of matching GISAID samples ranged from 5.8% to 8.2%. Overall, a significant fraction of patients (13/30) had viral mutation signatures which were relatively unique, even when using this relaxed criterion.

We further evaluated the frequency of specific genetic fingerprints. The P125 viral sample matched the most common GISAID fingerprint consisting of 3037C>T and 23403A>G **(Figure 3B)**, indicative of a common viral strain. The P155 viral sample matched the 3rd most common fingerprint in which there are no variants distinct from the NC_045512.2 reference which originated from Wuhan. The P125 viral sample had a common mutation signature with 17,851 exact matches **(Figure 8B)**, and P155 has 1,587 exact matches. Seven viral samples did not match any of the top 20 most common strain fingerprints. This included viral samples from P128, P133, P137, P139, P140, P146 and P152 which had a range of 1 to 66 matches. These samples had relatively unique fingerprints given that they were present in less than 0.001% in the larger GISAID data set. Overall, our results suggest that unique and relatively rare viral mutation fingerprints can be identified among a significant number of patients.

## DISCUSSION

With the rapid transmission of SARS-CoV-2 around the world, different strains have emerged as defined by the propagation of viruses with new mutations. Our study leveraged a systematic k-mer based analysis of the SARS-CoV-2 to identify critical regions of viral sequence conservation and provide mutation indexes across tens of thousands of patients. We used this sequence analysis for the development of a robust deep sequencing assay to detect viral mutations with high sensitivity. Then, we compared mutation profiles from a clinical sample set with those reported in GISAID. Based on the results of our study, this approach has the possibility of providing a highly scalable and integrated framework for identifying viral genetic fingerprints among patients that are relatively unique. Importantly, this process may facilitate molecular contract tracing where one compares viral mutation fingerprints among infected individuals.

The increasing availability of SARS-CoV-2 genome sequences provides biomedical and clinical researchers with a rich resource for investigating pandemic spread. However, traditional alignment-based analysis techniques lead to challenges when applied on a scale of thousands of genome sequences. Our analysis is independent from coordinate systems of any given genome, and is therefore independent of the choice of a specific SARS-CoV-2 reference. Comparisons of viral genome assemblies scale exponentially as the number of samples increases. However, our k-mer counting approach for identifying novel pangenome features scales linearly and therefore is computationally efficient. This approach enabled us to rapidly survey viral pangenome features such as genome conservation and mutation signatures from thousands of viral samples.

During the course of the SARS epidemic in 2003-2004, a number of mutations and deletions in *orf8* became predominant among the infected population [19-21]. A 29-nucleotide deletion in SARS-CoV *orf8* was observed in nearly all cases diagnosed during the middle and end of the outbreak, and complete or almost complete deletions of *orf8* were observed at the end of the epidemic [19-21]. Evidence from our pangenome study suggests that SARS-CoV-2 *orf8* shows evidence of evolutionary divergence with less than 10% of all bases sharing an anchor 25-mer sequence **(Figure 2)**. Notably, we observed six total mutations between *orf7* and *orf8* in our clinical sample set. Three were nonsynonymous mutations (27874 C > T in samples P144 and P146, 27925 C > T in sample P138, 27970 C > T in P132) and are expected to result in a change from threonine to isoleucine **(Supplementary Table 9)**. It will require additional studies to determine the consequences of these changes on viral fitness and disease.

Currently, there are a number of methods for detecting COVID-19 [22-24]. Quantitative reverse transcription polymerase chain reaction **(RT-qPCR)** is the most widely used SARS-CoV-2 diagnostic test, and is considered the ‘gold standard’ for detection due to its high specificity. [23] RT-qPCR diagnostic tests are based on the detection of nucleic acid from SARS-CoV-2 in respiratory specimens (such as nasopharyngeal and oropharyngeal swabs, sputum, and bronchoalveolar lavage fluid) collected from individuals suspected of COVID-19 infection. Serological antibody tests detect IgM and IgG generated in response to SARS-CoV-2 from blood samples. Antibody tests can provide information regarding current and previous SARS-CoV-2 infection, as well as potential immunity [25]. These molecular tests do not provide information about signatures of viral mutations, emergence of new strains or viral fingerprint patterns that are associated with transmission. Thus, sequencing of viral samples has an important role in diagnostic testing and disease control.

For our study, we developed a robust deep sequencing assay to ascertain quasispecies mutations and generate mutation signatures. We targeted 40% of the SARS-CoV-2 genome using primers from highly conserved regions based on our pangenome analysis. In the future, these same conserved primers can be used to detect novel mutations. By virtue of designing primers in conserved regions, we expect that our assay will be robust in performance, where we expect the primers to still bind to conserved regions of interest in SARS-CoV-2. Notably, when performing the same type of k-mer analysis to primer sets used in the ARTIC network’s amplicon sequencing assay, we observed that only 57% of the primers appeared in at least 99% of the 75,681k GISAID dataset. This indicates that natural variations observed in SARS-CoV-2 may impact the performance of primer binding as well as obscure mutations by imperfect primer annealing during PCR.

We restricted the number of amplicons to improve the multiplexed amplification of viral genome sequences. However, one limitation of this deep sequencing approach is that the coverage is restricted to only approximately 40% of the viral genome. As a result, not all mutations will be detected. To overcome this issue, future iterations of assay design will expand the number of amplicons for broader coverage, while maintaining primers in conserved sequences. Thus, based on our pangenome analysis, there are opportunities to target mutation prone sequences as they appear in the population.

Our sequencing assay possesses important operational advantages compared to other molecular detection methods. Following sample processing, numerous individual specimens can be pooled during library preparation and maintain their unique identity. Identification of individual samples relies on assignments from DNA-based sample barcodes. Another advantage is the use of a library normalization procedure that eliminates the need for library balancing and simplifies the workflow for sequencing. For small genomes like SARS-CoV-2, one can sequence tens of thousands of samples in single sequencing run, depending on the capacity of the sequencer. This scalability feature makes the analysis of large numbers of samples feasible compared to other assays which require that samples be maintained in individual wells. The operational scalability of NGS also enables one to conduct large scale population screening with the potential for significant cost reduction compared to other methods.

## CONCLUSIONS

Overall, the characterization of SARS-CoV-2 genetic variation provides insight into the paths of transmission and selection processes that may influence infection rates. Thus, viral mutations provide genetic fingerprints that cover a range of frequencies, such as the dominant species commonly observed among infected populations to less prevalent quasispecies that are limited to an individual or small number of patients. Analysis of lower frequency viral fingerprints may provide a way of conducting molecular contact tracing.

## Data Availability

Sequence data is available at the National Institutes of Health's Sequence Read Archive under BioProject Accession ID of PRJNA663917.

## ABBREVIATIONS

Bp: base pair
CDC: Centers for Disease Control and Prevention
cDNA: complementary DNA
EUA: Emergency Use Authorization
FDA: Food and Drug Administration
GISAID: Global Initiative on Sharing All Influenza Data
gnomAD: Genome Aggregation Database
kb: kilobase
NCBI: National Center for Biotechnology Information
NGS: next-generation sequencing
nsp: non-structural protein
nt: nucleotide
ORF: open reading frame
RdRp: RNA-dependent RNA polymerase
RT: reverse transcription
ViPR: Virus Pathogen Resource

## METHODS

### Viral samples

This study was conducted in compliance with the Helsinki Declaration. The Institutional Review Board at Stanford University School of Medicine approved the study protocol (IRB-56088). A total of 100 extracted RNA specimens from clinical nasopharyngeal swabs were obtained from the Stanford University Clinical Virology Lab (Stanford University, Stanford, CA). The specimens consisted of 30 positives and 70 negatives externally confirmed by an Emergency Use Authorization (EUA) approved SARS-CoV-2 detection RT-qPCR assay (FDA EUA200036). All samples were fully anonymous with no identifiers.

### Saliva sample collection and processing

Saliva samples were obtained from healthy donors with no symptoms of respiratory infection (presumed SARS-CoV-2 negative) using OMNIgene ORAL saliva collection tubes (DNA Genotek, Ottawa, CA, catalog no. OM-505). Cells were lysed using the buffer provided in the caps of the sample collection tubes. Lysed saliva samples were inactivated at 65°C for 15 minutes prior to RNA extraction. We extracted RNA using the Promega Maxwell Viral Total Nucleic Acid Kit (Promega, Madison, WI, catalog no. 639209) according to manufacturer instructions. RNA samples were eluted with 40 µL of RNAse-free water. Human RNA extracted from saliva samples of SARS-CoV-2 negative donors was quantified using the Qubit RNA HS Assay Kit (Thermo Fisher Scientific, catalog no. Q32852) and stored at -80°C.

### Genome sequences of SARS-CoV-2, other human viruses and bacteria

We accessed the data available from three different sources: SARS-CoV-2 genomes from Global Initiative on Sharing All Influenza Data **(GISAID)**; other human coronaviruses from Virus Pathogen Resource (**ViPR**); and lastly other human viruses and bacteria sequences from the National Center for Biotechnology Information **(NCBI) (Additional file 1; Supplementary Table 1)**. According to GISAID, the definition of “complete” means the genome is greater than 2,900 bp and “high-coverage” means only select entries with 1% N’s and <0.05% unique amino acid mutations (not seen in other sequences in database) and no insertion/deletion unless verified by submitter. We also identified 447 human coronavirus genomes by selecting “host: human”, “complete genome”, and “date: up to 2019 Oct” from ViPR [26]. Furthermore, we obtained 804 virus genomes with human host from NCBI viral genomes [27].

We identified and removed coronaviruses from the compiled 804 virus genomes. As a result, we had our own list of the virus genomes consisting of 42 influenza viruses, and 320 human viruses. “Respiratory syncytial virus” was added to our human virus list in acknowledgement of the existence of this virus in the FDA’s cross-reactivity list. With this, all viruses on the FDA’s cross-reactivity list were included in our list. Additionally, we retrieved and added the reference genomes of 27 bacteria in the FDA’s cross-reactivity list. We used *Rothia mucilaginosa* DY-18 searched by “*Stomatococcus mucilaginosus,”* which is the new name of *Staphylococcus salivarius*. NCBI selected *Pneumocystis jirovecii* RU7 (assembly Pneu_jiro_RU7_V2) as reference genomes for *Pneumocystis jirovecii* (PJP).

### Metadata of 25-mers present in SARS-CoV-2 genomes and other genomes of interest

We generated a searchable index of location metadata for all 25-mers found in 3,968 SARS-CoV-2 genomes using our in-house computational tool. We also included all 25-mers in other 837 human viruses and bacteria and human genomes, GRCh38 **(Additional file 1; Supplementary Table 1)**. The metadata provides the locations of all distinct canonical k-mers in all sequences of interest, such as the 3,968 SARS-CoV-2 genomes included in this analysis. A canonical k-mer is the lexicographically smaller sequence of a k-mer and its reverse complement. For instance, the metadata for the 25-mer ‘AGGGACTATTCCCACCCAAGAATAG’ in the SARS-CoV-2 genomes contains the following information: sequence ID, position, and strand, appearing in three sequences (distinct entries separated by semicolons):

*USA/CruiseA14/EPI_ISL_413619/2020-02-25, 11742-11766*,+;

*Shangrao/JX1974/EPI_ISL_421258/2020-02-08,11718-11742,* +;

*USA/TX_2020/EPI_ISL_419561/2020-02-29,11742-11766,+;*

Our in-house computational tool allows for fast generation of an index associating k-mers with arbitrary metadata, and enables queries of this index that are either exact or approximate, permitting some number of mismatching base pairs in a k-mer. This tool is written in the Julia programming language, a high-performance, general programming language interoperable with Python, R, C, and Fortran; we also provide a command-line interface for language-agnostic use and to support use in pipelines with other software. Our analysis was run on an AMD EPYC 7501 32-Core Processor, 503 GiB, Linux version 4.4.0-184-generic #214-Ubuntu SMP.

Intuitively, a k-mer is “specific” to a “target” set of genomes with respect to another, “off-target” set of genomes if the k-mer occurs in many of the target genomes and does not occur in any of the off-target genomes, even allowing for a few mismatching base pairs in the off-target set. Moreover, a k-mer is “unique” within a genome if it occurs exactly once within that genome. A k-mer is “X% conserved” within a set of genomes if it occurs in at least X% of those genomes. “Anchor” k-mers are both 100% conserved and unique within each genome. A k-mer is “X% specific with up to M mismatches” to a target set of genomes G with respect to off-target set of genomes G’ if it is X% conserved in G, and does not occur in any genome in G’ even if we allow up to M mismatching base pairs.

In the example above, a 25-mer appears in three SARS-CoV-2 genomes at the coordinates printed. These coordinates are relative to the respective sample genomes. Crucially, our method supports fast, approximate search, necessary to satisfy constraints of maximum 80% homology to other human viruses/bacteria and 90% homology to GRCh38 as outlined by the FDA’s SARS-CoV-2 detection assay EUA criteria. Our 25-mer index stores location metadata for all 25-mers in a list of genomes, and also allows approximate lookup by allowing up to M mismatching base pairs when querying the set of locations at which a 25-mer appears. For example, the 25-mer AATTGTACTGTTTTTAACAAAGCTT is conserved and unique among the 3,968 SARS-CoV-2 sequences, but is not specific to SARS-CoV-2 because it occurs within 2 mismatches at 2 locations in GRCh38 (see below; dots indicate base pairs matching GRCh38, and capital letters denote mismatches).

*‘seq’: ‘*………*T*…….*A*…….*’,’chr1’, ‘pos’:’183137629-183137653’, ‘strand’:’-’*,

*‘seq’: ‘*….*A*………*C’, ‘chr16’, ‘pos’:’457072-457096’, ‘strand’:’-’*

This enables us to identify 25-mers that have no approximate matches (1) up to 4 mismatched base pairs with the genomes of all human coronaviruses, other human viruses, influenza, and the 27 bacteria included in the FDA’s SARS-CoV-2 detection assay cross-reactivity list, and (2) up to 2 mismatched base pairs with the human genome (GRCh38). This enables us to identify 25-mers that have no approximate matches (1) up to 4 mismatched base pairs with the genomes of all human coronaviruses, other human viruses, influenza, and the 27 bacteria included in the FDA’s SARS-CoV-2 detection assay cross-reactivity list, and (2) up to 2 mismatched base pairs with the human genome (GRCh38).

We also constructed a k-mer index containing all k-mers induced by all possible single substitution mutations, two reported deletions, and multi-nucleotides substitutions in the N gene. To make this index, for each variation we took the set of k-mers from the SARS-CoV-2 reference genome that overlaps the reference allele position – the “reference set”, modified these k-mers to instead contain the alternate allele – the “alternate set”, and then associated each of these modified k-mers with metadata describing the variation. For example, suppose that (1) k=3, (2) our variant is the substitution from A to C at position 1000, and (3) the reference sequence from position 998 to 1002 is CGATT. Then the reference set of 3-mers is (CG**A**, G**A**T, **A**TT) (reference allele in bold), the alternate set of 3-mers is (CG**T**, G**T**T, T**T**T) (alternate allele in bold), and our k-mer index would map each of the alternate 3-mers to a description of this variant. An “alternate” k-mer can be associated with multiple distinct mutations, but for k=25 we found that most of the alternate 25-mers were associated uniquely with exactly one mutation. Any 25-mers that were not associated with exactly one mutation removed from the index. This 25-mer index of mutation information allows us to survey efficiently mutations in viral assemblies from GISAID without using a sequence alignment. In other words, we can get the frequency of a particular mutation simply by counting the number of genomes that have the mutation-specific 25-mers derived from the mutation.

### Targeted sequencing primers

We generated a list of candidate primer pairs for targeted sequencing. Let G_cov2 denote the set of 3,968 SARS-CoV-2 genomes, let G_other denote the set of viral and bacterial genomes, and let G_human be the human reference genome GRCh38. A candidate primer pair consists of two 25-mers denoted x (forward primer) and y (reverse primer) that satisfy four properties: conservation and uniqueness; specificity; positional constraints; and compositional constraints. To be conserved and unique, both x and y must appear in all 3,968 SARS-CoV-2 sequences exactly once. In order to be specific, at least x or y in a pair must not occur in G_other, even allowing for up to 4 out of 25 mismatched base pairs (i.e. maximum 80% homology to any 25- mer in G_cov2). Additionally, x or y must not occur in G_human even allowing for up to 2 out of 25 mismatched base pairs. If the primer pair meets the requirements for positional constraints, the maximum distance between x and y in any sequence in G_cov2 must be 2,500 base pairs at the most, not including the combined 50 base pairs of x and y. Finally, the compositional constraints mean that the GC content of x and y may differ by 2 out of 25 base pairs at a maximum.

We identified 67,478 primer pairs that met the above criteria. In addition, we selected *RPP30* as a human control, which was used in the Centers for Disease Control and Prevention’s (CDC) RT-PCR diagnostic testing. To ensure all copies of *RPP30* were amplified, we designed primers for both human cDNA and genomic DNA. We selected highly conserved sequences such that genetic variation among individuals would not affect the ability of the primers to anneal to their complement. Specifically, we selected conserved regions that flank regions of high variability, according to frequencies in the Genome Aggregation Database **(gnomAD)**. For human DNA, we chose two forward and reverse primer pairs. The first primer pair sequences *RPP30* across exon 1 to produce an amplicon of 0.476kb. The second primer pair sequences a region between exons 6 and 7 to produce an amplicon of 1.277kb. Within the amplicon of the second primer pair, there are three regions with high variability. If the variability of each region is 50% such that one out of two individuals is likely to have a single nucleotide polymorphism, then there are 8 unique amplicons that can arise from that primer pair. Each individual therefore has a slightly distinct amplicon that can be used to differentiate patient samples within a pool.

### Targeted sequencing of SARS-CoV-2

We first experimentally confirmed that the saliva samples were negative for SARS-CoV-2 using a commercial one-step RT-qPCR assay (Thermo Fisher Scientific, catalog no. A15299) on the StepOnePlus Real Time Quantitative PCR instrument (Thermo Fisher Scientific, Applied Biosystems). Briefly, RNA samples were reverse transcribed to cDNA and then subjected to 45 cycles of quantitative PCR according to the following recommended conditions: UNG incubation at 25°C for 2 minutes, reverse transcription incubation at 50°C for 15 minutes, enzyme activation at 95°C for 2 minutes, 45 cycles of amplification consisting of denaturation at 95°C for 3 seconds followed by annealing/extension at 60°C for 30 seconds, and a final infinite holding step at 4°C. We used previously described primers and 6-carboxyfluorescein (FAM)-labeled hydrolysis probes targeting three regions of the SARS-CoV-2 nucleocapsid protein (N) gene. We also used additional primer probe set targeted a human housekeeping (*RPP30*) gene as an internal/extraction control. The primers and probes used for both the SARS-CoV-2 targets and the human *RPP30* targets were identical to those described in the CDC nCoV-19 assay. No amplification for SARS-CoV-2 targets occurred in any of the negative control samples.

We developed a two-step multiplex PCR protocol for the reverse transcription and PCR amplification of SARS-CoV-2 targets from both the clinical and contrived samples. Briefly, cDNA was synthesized from RNA samples using random hexamer priming and ProtoScript II First Strand cDNA Synthesis Kit (New England Biolabs, catalog no. E6560L) according to manufacturer instructions. We generated amplicons from cDNA samples using the Titanium Taq PCR Kit (TaKaRa Bio, catalog no. 639210) according to the following recommended cycling conditions: enzyme activation at 95°C for 1 minute, 40 cycles of denaturation at 95°C for 30 seconds followed by annealing/extension at 68°C for 2.5 minutes, final extension at 68°C for 3 minutes, and a final infinite holding step at 4°C. One microliter of cDNA from contrived samples was used for the PCR reaction, and five microliters was used for the patient samples to maximize the amount of genomic material to be amplified. We used a panel of six SARS-CoV-2- specific primers sets targeting non-overlapping regions ranging from 1 kb to 2.5 kb in length across the viral genome, and a primer set targeted the human *RPP30* gene as an extraction control. PCR was performed on a Veriti Thermal Cycler (Thermo Fisher Scientific, Applied Biosystems).

We performed massively parallel library preparation using a pooled tagmentation approach called plexWell (Seqwell, catalog no. PW384). Briefly, this library preparation consists of a two-step transposase-based process referred to as tagmentation, where sequencing adapters are randomly inserted into the PCR amplicons DNA by transposition. The first tagmentation step incorporates well-specific Illumina i7-Read 2 barcodes. After pooling, a second tagmentation step incorporating a plate-specific Illumina i5-Read 1 barcode finishes the library. The tagmentation enzyme is reagent-limiting in each well, which obviates the need for individual sample normalization. After PCR amplification, we performed a 1:10 dilution of the PCR plate for use with the plexWell protocol per manufacturer instructions, with the only adjustment being a final magnetic bead cleanup using a ratio of 0.9X beads rather than 0.75X to enrich for shorter fragments. After library preparation, the sample was quantified by Qubit (Thermo Fisher Scientific, catalog no. Q32851), visualized on an 2% E-Gel EX cartridge (Thermo Fisher Scientific, catalog no. G402002), and loaded on an iSeq 100 cartridge (Illumina, catalog no. 20021533) with 5% PhiX library control v3 (Illumina, catalog no. FC-110-3001) spike-in with 151bp paired-end reads and 8bp dual indexing.

### Digital PCR

We measured the concentrations of both human (*RPP30*) cDNA and SARS-CoV-2 cDNA in the contrived samples by performing multiplexed droplet digital PCR (ddPCR) using ddPCR Supermix for Probes (no dUTP) (Bio-Rad, catalog no. 1863024), the CDC nCoV-19 N1 assay (IDT, catalog no. 10006606), and a commercially available human *RPP30* genomic DNA assay (Bio-Rad, catalog no. 10042961) on the QX200 Droplet Digital PCR System (Bio-Rad). The N1 assay included a 6-carboxyfluorescein (FAM)-labeled hydrolysis probe and the *RPP30* assay included a hexachlorofluorescein (HEX)-labeled hydrolysis probe. Viral and human genomic targets were amplified following droplet generation according to the following recommended cycling conditions: incubation at 25°C for 3 minutes, enzyme activation at 95°C for 10 minutes, 40 cycles of denaturation at 94°C for 30 seconds followed by annealing/extension at 60°C for 1 minute, enzyme deactivation at 98°C for 10 minutes, and a final infinite holding step at 4°C. Following amplification, droplets were analyzed using the QX200 Droplet Reader and QuantaSoft Software (Bio-Rad).

### Control samples

We contrived positive samples by spiking genomic RNA from SARS-CoV-2 (ATCC, Genomic RNA from SARS-Related Coronavirus 2, Isolate USA-WA1/2020, catalog no. NR-52285) into the PCR-confirmed SARS-CoV-2 negative saliva RNA samples. To assess the limit of detection, we generated a series of 12 contrived samples at total inputs ranging from 1×10^3^ to 0 genome copies. We generated an additional set of 3 SARS-CoV-2 dilutions at the lowest inputs (e.g. 2 copies, 1 copy, and 0 copies) for downstream cDNA synthesis to confirm the limit of detection for assay.

### Admixture analysis for detection of sub-clonal mutation allelic fraction

We created an additional set of 12 contrived samples by spiking genomic RNA isolates from two previously characterized SARS-CoV-2 strains (ATCC, Genomic RNA from SARS-Related Coronavirus 2, Isolate USA-WA1/2020 and ATCC, Genomic RNA from SARS-Related Coronavirus 2, Isolate Hong Kong/VM20001061/2020) into the same PCR-confirmed SARS-CoV-2 negative human RNA samples at different relative fractions **(Supplemental Table 2)**. All samples had the same total concentration of viral RNA (1,000 copies).

### Bioinformatic sequencing analysis

Raw sequence data underwent base calling and demultiplexing using bcl2fastq (v2.20). Reads were aligned using bwa (mem algorithm; v0.7.17) and processed into bam files using samtools (v1.10). As a reference genome, we generated a merged reference by concatenating GRCh38 and the ancestral SARS-CoV-2 reference sequence (NC_045512.2) into a single FASTA file and creating a new bwa reference. Reads aligning to either the human or viral genome were counted by using the command “samtools idxstats,” and per-base coverage metrics were analyzed using bedtools (v2.29) using the “bedtools coverage -d” command, selecting only for the SARS-CoV-2 genome.

For identification of variants in clinical samples, we performed variant calling using Sentieon (v201808.08) with the reference genome of SARS-CoV-2 (NC_045512.2) and the reference human genome (GRCh38). First, we preprocessed FASTQ files containing raw sequence data with quality control using the default setting of fastp. Second, the paired-end alignments from the filtered reads against were implemented by the Sentieon bwa binary. Next, deduplication and realignment around indels were performed on each sample: “sentieon driver --algo Dedup and --algo Realigner”. Fourth, we used the haplotype caller setting to conduct variant calling with base quality recalibration, which was derived from the recalibrated bam files: “sentieon driver --algo Haplotyper –annotation QD,MQ, MQRankSum,ReadPosRankSum,FS, SOR,DP”. Finally, variants consistent between two or more replicates from each respective patient sample were considered real mutations for each sample and were included in our downstream mutation profile analyses. Downstream analysis used R for generating plots and statistical calculations.

## DECLARATIONS

### Ethics approval and consent to participate

This study was conducted in compliance with the Helsinki Declaration. The Institutional Review Board (IRB) at Stanford University School of Medicine approved the study protocol (IRB-56088).

### Consent for publication

Not applicable.

### Availability of data and material

Sequence data is available at the National Institutes of Health’s Sequence Read Archive **(SRA)** under BioProject Accession ID of PRJNA663917.

### Authors’ contributions

HJ.L., B.T.L. and H.P.J. conceived and designed the study; HJ.L., and D.P. implemented the computational method. B.A.P. provided viral samples and quantitative PCR results. D.P. and HJ.L. provided data. B.T.L, A.C.H. and G.S. conducted the sequencing experiments. A.C.H. and J.C. conducted molecular genetic analysis. A.A. and A.C.H. supported the translational component of the study. HJ.L., D.P., B.T.L., G.S. and H.P.J. analyzed the data. All authors contributed to the manuscript writing. H.P.J. supervised the project.

## Acknowledgements

We would like to thank Anuja Sathe, Carlos Ayala, and Maddy McNamara for their feedback on the manuscript as well as Stephanie Greer and Susan Grimes for their assistance with variant calling. We would also like to acknowledge Joe Mellor and Danielle Bodnar at Seqwell for extensive technical discussions. Finally, we acknowledge that figures 1, 3 and 7 were created with BioRender.com.

## ADDITIONAL FILES

**Additional file 1**: Supplementary Table S1 – S9. Format: PDF

**Additional file 2:** The GISAID identifies of 3,968 and 75,681 SARS-CoV-2 genome assemblies. Format: XLSX

**Additional file 3:** The mutation landscape of 4K and 75K SARS-CoV-2 genomes. Format: XLSX

**Additional file 4:** Sequencing coverage metrics. Format: XLSX

