## Supplemental Materials for "Profiling SARS-CoV-2 mutation fingerprints that range from the viral pangenome to individual infection quasispecies"

**TITLE**

**Institutions**

[1]Division of Oncology, Department of Medicine, Stanford University School of Medicine, Stanford, CA, 94305, United States

[2]Stanford Genome Technology Center West, Stanford University, Palo Alto, CA, 94304, United States

[3]Department of Pathology, Stanford University School of Medicine, Stanford, CA, 94305, United States

[4]Department of Medicine, Division of Infectious Diseases and Geographic Medicine, Stanford University School of Medicine, Stanford, CA, 94305, United States

**Corresponding authors**

Hanlee P. Ji

HoJoon Lee

Division of Oncology, Department of Medicine – Stanford University School of Medicine

269 Campus Drive, CCSR 1120, Stanford, CA 94305-5151

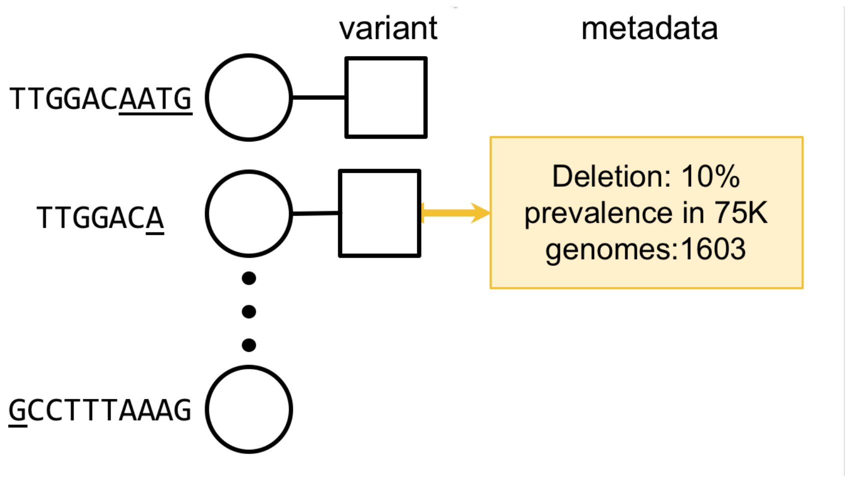


**Supplementary Figure 1. Metadata kmer indexing for mutations.** We incorporated metadata annotating mutation features from the k-mer derived from individual SARS-CoV-2 genomes present in GISAID. This metadata includes the nature of the mutation and its frequency among the sample set.

**SUPPLEMENTARY TABLES**

**Supplementary Table 1.** Genome assemblies included pangenome k-mer study.

| **Organism or virus** | **Number of unique genome assemblies** |
|---|---|
| SARS-CoV-2 | 3,968 |
| Bacterial genomes | 89 |
| GRCh38 | 1 |
| Influenza genomes | 42 |
| Other human-host viruses | 321 |
| Other human-host coronaviruses | 447 |

**Supplementary Table 2.** K-mer filter criteria for primer design and selection.

| **Filter criterion** | **# 21-mers** | **# 23-mers** | **# 25-mers** | **# 27-mers** | **# 29-mers** |
|---|---|---|---|---|---|
| All k-mers in 3,968 SARS-CoV-2 genomes | 84276 | 89348 | 94402 | 99446 | 104488 |
| and conserved and unique in all SARS-CoV-2 genomes | 2777 | 2347 | 1977 | 1664 | 1406 |
| and max k-5 match to 89 bacteria genomes | 1 | 402 | 1408 | 1584 | 1397 |
| and max k-3 bp match to human genome (GRCh38) | 0 | 289 | 1336 | 1575 | 1397 |
| and max k-5 bp match to 42 influenza genomes | 0 | 289 | 1336 | 1575 | 1397 |
| and max k-5 bp match to 321 other human viruses | 0 | 259 | 1313 | 1573 | 1397 |
| and max k-5 bp match to 447 other human coronaviruses | 0 | 63 | 433 | 637 | 642 |

**Supplementary Table 3.** Accession numbers of genomes included in *in silico* cross-reactivity analyses.

| **Bacteria/Virus** | **Accession Numbers** |
|---|---|
| *Chlamydia pneumoniae* | CP001713.1, AE009440.1, AE009440.1 |
| *Haemophilus influenza* | CP031689.1, NC_000907.1 |
| *Legionella pneumophila* | CP041668.1, CP025491.2 |
| *Mycobacterium tuberculosis* | CP000717.1 |
| *Streptococcus pneumoniae* | CP007593.1, CP001845.1 |
| *Streptococcus pyogenes* | AE009949.1 |
| *Pneumocystis jirovecii* | AY685194.1, AY127566.1, AY130996.1, JX499143.1 |
| *Candida albicans* | NC_002653.1, NC_002653.1, NC_002653.1 |
| *Pseudomonas aeruginosa* | NZ_CP040684.1, NZ_CP027174.1, NZ_CP007147.1 |
| *Staphylococcus epidermidis* | NZ_CP018842.1 |
| *Streptococcus salivarius* | NZ_CP040804.1, NZ_CP018187.1, NZ_CP018189.1, NZ_CP020451.2, NZ_CP020451.2 |
| Human metapneumovirus | KJ627397.1, AY525843.1, KJ627383.1, AF371337.2 |
| Parainfluenza virus 1 | M14887.1, AF457102.1, KF687307.1, AF457102.1, KX639498.1 |
| Parainfluenza virus 2 | NC_003443.1, AF533011.1, KM190939.1 |
| Parainfluenza virus 3 | KM190938.1, KY973556.1, MH678682.1 |
| Parainfluenza virus 4 | NC_021928.1, MH892407.1, KY460515.1, KF483663.1 |
| Influenza A virus | AB827993.1, AB818499.1, NC_007367.1, HE589468.1, AB822988.1, NC_007371.1 |
| Influenza B virus | NC_002206.1, NC_002211.1, NC_002205.1, NC_002207.1, NC_002205.1, NC_002211.1 |
| Enterovirus | KP202389.1, MK593172.1, FJ445142.1, FJ445125.1, AB647318.1 |
| Rhinovirus | MG950178.1, DQ473499.1, FJ445142.1, FJ445174.1, FJ445125.1, FJ445147.1 |

**Supplementary Table 4.** Multiplexed PCR primers used for amplicon generation and subsequent Illumina sequencing.

| | | Forward primers | | | Reverse primers | | | |
|---|---|---|---|---|---|---|---|---|
| Primer pair # | Target | Genomic coordinates | | Sequence | Genomic coordinates | | Sequence | Amplicon length (kb) |
| | | Start | End | | Start | End | | |
| 1 | SARS-CoV-2 | 1821 | 1845 | GTGCCTGGAATATTGGTGAACAGAA | 3048 | 3072 | CAATCACCTTCTTCTTCATCCTCAT | 1.252 |
| 3 | SARS-CoV-2 | 7450 | 7474 | AAGTTATGTGCATGTTGTAGACGGT | 8495 | 8519 | TAACAACTTGTCTAGTAGTTGCACA | 1.070 |
| 4 | SARS-CoV-2 | 9971 | 9995 | AAGGCTCTCAATGACTTCAGTAACT | 12446 | 12470 | TGGCTGCTGTTGTAAGAGGTATTAT | 2.500 |
| 6 | SARS-CoV-2 | 16264 | 16288 | TCACAGACTTCATTAAGATGTGGTG | 18267 | 18291 | ACGTACATGTCTTATAGCTTCTTCG | 2.028 |
| 7 | SARS-CoV-2 | 20996 | 21020 | GTGATTGTGCAACTGTACATACAGC | 23638 | 23662 | ACCAAGTGACATAGTGTAGGCAATG | 2.667 |
| 8 | SARS-CoV-2 | 26098 | 26122 | ATTGTTGATGAGCCTGAAGAACATG | 28480 | 28504 | ATTGGTGTTAATTGGAACGCCTTGT | 2.407 |
| 2 | human *RPP30* | 90889888 | 90889912 | CTTGTCATCGCATTTCTGTCATGTG | 90891190 | 90891214 | AGGTGGTCCTATAGATTTCAGAGGG | 1.327 |

**Supplementary Table 5.** Concentrations of serially diluted contrived SARS-CoV-2 samples used for analytical sensitivity and specificity testing.

| Sample # | SARS-CoV-2 concentration (copies/µl) |
|---|---|
| **1** | 1.0000E+03 |
| **2** | 5.0000E+02 |
| **3** | 2.5000E+02 |
| **4** | 1.2500E+02 |
| **5** | 6.2500E+01 |
| **6** | 3.1250E+01 |
| **7** | 1.5625E+01 |
| **8** | 7.8125E+00 |
| **9** | 3.9063E+00 |
| **10** | 1.9531E+00 |
| **11** | 9.7656E-01 |
| **12** | 0.0000E+00 |

**Supplementary Table 6.** Strain-specific relative fractions of admixed SARS-CoV-2 contrived samples used for validation of k-mer based analysis.

| Sample # | Admixture Fraction | |
|---|---|---|
| | USA-WA1/2020 strain | Hong Kong/VM2000106 1/2020 strain |
| 1 | 1.00 | 0.00 |
| 2 | 0.99 | 0.01 |
| 3 | 0.95 | 0.05 |
| 4 | 0.90 | 0.10 |
| 5 | 0.75 | 0.25 |
| 6 | 0.50 | 0.50 |
| 7 | 0.25 | 0.75 |
| 8 | 0.10 | 0.90 |
| 9 | 0.05 | 0.95 |
| 10 | 0.01 | 0.99 |
| 11 | 0.00 | 1.00 |
| 12 | 0.00 | 0.00 |

**Supplementary Table 7.** Strain-specific mutations detected from admixed SARS-CoV-2 contrived samples.

| Reference position NC_045512.2 | Reference base | Strain specific mutations | |
|---|---|---|---|
| | | Hong Kong/VM20001061/2020 | USA-WA1/2020 |
| 18060 | C | | T |
| 21636 | C | T | |
| 22661 | G | T | |
| 23607 | G | A | |
| 26729 | T | C | |
| 27266-27292 | | DEL | |
| 28077 | G | C | |

**Supplementary Table 8**. Cycle threshold ($C_T$) values of externally tested clinical SARS-CoV-2 samples used for sequencing.

| Number | Viral Sample ID | CT Value SARS-CoV-2 qPCR |
|---|---|---|
| 1 | 152 | 15.36 |
| 2 | 144 | 16.09 |
| 3 | 146 | 16.37 |
| 4 | 125 | 19.19 |
| 5 | 140 | 19.48 |
| 6 | 137 | 19.58 |
| 7 | 132 | 20.58 |
| 8 | 145 | 20.94 |
| 9 | 155 | 21.08 |
| 10 | 133 | 21.14 |
| 11 | 128 | 21.45 |
| 12 | 151 | 22.19 |
| 13 | 142 | 23.07 |
| 14 | 139 | 23.41 |
| 15 | 143 | 27.16 |
| 16 | 126 | 28.16 |
| 17 | 138 | 28.54 |
| 18 | 131 | 28.71 |
| 19 | 122 | 29.08 |
| 20 | 162 | 29.79 |
| 21 | 124 | 31.30 |
| 22 | 127 | 32.80 |
| 23 | 121 | 34.30 |
| 24 | 129 | 34.90 |
| 25 | 130 | 35.50 |
| 26 | 149 | 35.93 |
| 27 | 147 | 37.31 |
| 28 | 123 | 38.20 |
| 29 | 150 | 38.35 |
| 30 | 141 | 39.06 |

**Supplemental Table 9. Expected translational changes from mutations in ORF7a and ORF8 from clinical samples.**

| **Mutation** | 27641 C > T | 27670 G > T | 27874 C > T | 27925 C > T | 27970 C > T | 28144 T > C |
|---|---|---|---|---|---|---|
| **Translational change** | S83L | V93F | T40I | T11I | T26I | L84S |
| **Protein** | ORF7a | ORF7a | ORF7b | ORF8 | ORF8 | ORF8 |
| **Wild type AA** | serine | valine | threonine | threonine | threonine | leucine |
| **Wild type AA polarity** | polar | nonpolar hydrophobic | polar | polar | polar | nonpolar hydrophobic |
| **Wild type AA MW (Da)** | 105.09 | 117.15 | 119.12 | 119.12 | 119.12 | 131.18 |
| **Mutant AA** | leucine | phenylalanine | isoleucine | isoleucine | isoleucine | serine |
| **Mutant polarity** | nonpolar hydrophobic | nonpolar aromatic | nonpolar hydrophobic | nonpolar hydrophobic | nonpolar hydrophobic | polar |
| **Mutant MW (Da)** | 131.18 | 204.23 | 131.18 | 131.18 | 131.18 | 105.09 |
| **Change in polarity** | from polar to nonpolar hydrophobic | from nonpolar hydrophobic to nonpolar aromatic | from polar to nonpolar hydrophobic | from polar to nonpolar hydrophobic | from polar to nonpolar hydrophobic | from nonpolar hydrophobic to polar |
| **Change in MW (Da)** | 26.09 | 87.08 | 12.06 | 12.06 | 12.06 | -26.09 |
| **Samples** | Pos_126 | Pos_139 | Pos_144, Pos_146 | Pos_138 | Pos_132 | Pos_145, Ctrl_001 |
| **GISAID frequency** *(n=75,681)* | 31 | 35 | 40 | 22 | 66 | 4655 |
| **GISAID %** | 0.040961404 | 0.046246746 | 0.052853424 | 0.029069383 | 0.08720815 | 6.150817246 |
